# Proteomic Changes in the Human Cerebrovasculature in Alzheimer’s Disease and Related Tauopathies Linked to Peripheral Biomarkers in Plasma and Cerebrospinal Fluid

**DOI:** 10.1101/2024.01.10.24301099

**Authors:** Aleksandra M. Wojtas, Eric B. Dammer, Qi Guo, Lingyan Ping, Ananth Shantaraman, Duc M. Duong, Luming Yin, Edward J. Fox, Fatemeh Seifar, Edward B. Lee, Erik C. B. Johnson, James J. Lah, Allan I. Levey, Yona Levites, Srikant Rangaraju, Todd E. Golde, Nicholas T. Seyfried

## Abstract

Dysfunction of the neurovascular unit stands as a significant pathological hallmark of Alzheimer’s disease (AD) and age-related neurodegenerative diseases. Nevertheless, detecting vascular changes in the brain within bulk tissues has proven challenging, limiting our ability to characterize proteomic alterations from less abundant cell types. To address this challenge, we conducted quantitative proteomic analyses on both bulk brain tissues and cerebrovascular-enriched fractions from the same individuals, encompassing cognitively unimpaired control, progressive supranuclear palsy (PSP), and AD cases. Protein co-expression network analysis identified modules unique to the cerebrovascular fractions, specifically enriched with pericytes, endothelial cells, and smooth muscle cells. Many of these modules also exhibited significant correlations with amyloid plaques, cerebral amyloid angiopathy (CAA), and/or tau pathology in the brain. Notably, the protein products within AD genetic risk loci were found concentrated within modules unique to the vascular fractions, consistent with a role of cerebrovascular deficits in the etiology of AD. To prioritize peripheral AD biomarkers associated with vascular dysfunction, we assessed the overlap between differentially abundant proteins in AD cerebrospinal fluid (CSF) and plasma with a vascular-enriched network modules in the brain. This analysis highlighted matrisome proteins, SMOC1 and SMOC2, as being increased in CSF, plasma, and brain. Immunohistochemical analysis revealed SMOC1 deposition in both parenchymal plaques and CAA in the AD brain, whereas SMOC2 was predominantly localized to CAA. Collectively, these findings significantly enhance our understanding of the involvement of cerebrovascular abnormalities in AD, shedding light on potential biomarkers and molecular pathways associated with CAA and vascular dysfunction in neurodegenerative diseases.

## Introduction

Alzheimer’s disease (AD) is characterized by the accumulation of two core pathologies, amyloid beta (Aβ) plaques and phosphorylated tau neurofibrillary tangles (NFTs) in the brain, ultimately leading to dementia ^1-3^. However, mixed pathologies and comorbidities co-exist with Aβ plaques and tau NFTs in AD brains, including hippocampal sclerosis, Lewy body inclusions and cerebrovascular disease^4^. Notably, cerebrovascular dysfunction has been recognized as an early manifestation of AD contributing to cognitive decline and neurodegenerative changes in the brain^5,6^.

The cerebrovasculature forms a blood brain barrier (BBB) that exhibits low rates of transcytosis helping to protect the brain from potentially harmful peripheral factors, and which changes with age and in an accelerated manner in AD ^7,8^. Increased BBB permeability has been reported in the early stages of AD pathophysiology and has been hypothesized to play a central role in neurodegeneration^9-11^. In parallel to BBB breakdown, progressively reduced and dysregulated cerebral blood flow (CBF) and impaired hemodynamic responses are commonly seen in individuals with mild cognitive impairment or early AD. The exacerbated alterations of vasculature in AD have been in part linked to the accumulation of Aβ in brain parenchyma and cerebral blood vessels as cerebral amyloid angiopathy (CAA)^12^. Cerebrovascular deposits of Aβ are predominantly present in leptomeningeal and cortical arteries, arterioles, and veins (CAA type 2) but can also be found in capillaries (CAA type 1)^13^. CAA occurs in 85-95% of individuals with AD, contributing to higher risk for hemorrhages, infarcts, and ischemic lesions^12^. Although overall impact of CAA on rates of progression is typically modest, recent studies have suggested that the accumulation of CAA is accompanied by increased tau pathology^14,15^. Specifically, it has been proposed that severe CAA interacts with neuritic plaques leading to greater tau burden and more rapid cognitive decline^14^. Moreover, other types of amyloids, including Danish amyloid (ADan), also associated with blood vessels, have been shown to induce tau hyperphosphorylation and misfolding^16^. Functional changes in cerebrovasculature, including diminished CBF have also been correlated with tau pathology in primary tauopathies such as progressive supranuclear palsy (PSP) and frontotemporal dementia with parkinsonism-17 (FTDP-17) where amyloid deposition is not apparent^17-21^, further suggesting a dynamic interplay between cerebrovascular dysfunction and AD-related pathologies in accelerating disease progression. Currently, the precise impact of AD-related pathologies, including tau and CAA, on the cerebrovasculature at a molecular and cellular level is not fully understood.

Recent studies utilizing single-cell RNA-sequencing (scRNA-seq), have characterized the transcriptomes of human and mouse vascular cells in both healthy and disease contexts. These investigations have introduced the concept of transcriptional heterogeneity among vascular cells, delineating unique cellular states or subtypes based on their anatomical location and distinct functions^22-26^. These highly specialized structural, functional, and metabolic properties of endothelial cells, smooth muscle cells (SMCs), pericytes, and perivascular cells, termed the neurovascular unit (NVU), ensure optimal neuronal function. Moreover, it has been shown that vascular cells within the NVU exhibit unique age- and pathology-related transcriptional changes in human AD^23-26^. We and others have previously documented the discordance between transcriptomic and proteomic profiles of brain cells with transcriptomic alterations only modestly correlating with the proteomic changes in human AD brain^27-29^. Yet, the global proteomic systems-level architecture of brain vascular cells in the context of AD-related pathologies is still limited. Current proteomic analyses have overwhelmingly focused on bulk tissues^29-31^, restricting the ability to characterize proteomic alterations of less abundant cell types in the brain. Recent proteomic profiling of human cerebrovasculature has shown a remarkable overlap between the protein signature of CAA type 1 and the vascular proteome of cerebral autosomal-dominant arteriopathy with subcortical infarcts and leukoencephalopathy (CADASIL), but not observed as common to AD amyloid proteinopathy, suggesting a shared mechanism in various cerebral small vessel diseases^32^. However, it is still unclear whether the same proteomic changes are present in cases presenting with the more common form of CAA and whether other AD-related pathologies impact the cerebrovascular proteome. Thus, dissecting proteomes of vasculature in a disease setting will be a crucial step towards understanding the mechanisms underlying BBB breakdown, and pathological accumulation of amyloid in the cerebrovasculature.

Here, we implemented a tandem mass tag mass spectrometry (TMT-MS) based quantitative proteomics approach to comprehensively examine proteomic changes of human cerebrovasculature from postmortem brain tissues from AD, PSP, and non-demented control individuals (n=67 cases). The quantification of over 9,800 proteins allowed us to generate a deep protein co-expression network consisting of 93 modules or communities of proteins displaying wide biological heterogeneity associated with diverse brain pathologies. Importantly, the comparison of vascular proteome with proteomic data obtained from paired bulk brain tissues from the same individuals provided a unique opportunity to interrogate the relationship and preservation between the different fractions. We also investigated the enrichment of candidate gene products associated with the risk of AD or PSP identified in genome-wide association studies (GWAS) in protein modules within the cerebrovascular network. This uncovered several modules associated with endothelial, pericyte, and SMC biology. In particular, ‘Matrisome/Heparin binding’ module enriched with amyloid-associated proteins^33^ showed high specificity to AD with multiple module members altered in cases with pronounced vascular pathologies. Finally, by integrating the cerebrovascular network proteome with cerebrospinal fluid (CSF) and plasma proteomes from control and AD patients led to the identification of promising AD biomarkers enriched in the cerebrovasculature in brain. In summary, by taking advantage of our network-driven approach we nominated pathways strongly correlated with AD-related neuropathological traits and vascular biology that could provide the foundation for our understanding of the mechanisms governing AD-linked breakdown of BBB, dysregulation of the CBF, and pathological accumulation of protein aggregates in human vasculature.

## Results

### Comparative analysis of vascular and bulk proteomes in brain

Proteomic characterization of cerebrovasculature has been challenging owing to low abundance of vascular cells in the brain and inefficient capturing of vascular-specific proteins by bulk quantitative methods. Thus, to investigate the molecular definition of human brain vasculature in comparison to traditional bulk analysis, we isolated cerebral blood vessels from frozen frontal cortical tissues using density-mediated separation. To verify the enrichment of the cerebrovasculature in our preparations, we analyzed proteomic profiles of different brain fractions, including i) whole brain, ii) parenchyma/myelin, and iii) vascular fractions by label-free MS-based proteomics. We identified core clusters of highly enriched fraction-specific proteomic signatures. As expected, the cerebrovascular fraction was enriched in proteins specific to endothelial cells, SMCs, pericytes, and fibroblasts (**Supplemental Figure 1A**). Further validation of the enrichment of the human brain vasculature by Western blotting showed a significant increase in the levels of vascular specific proteins in the vascular fraction, including markers of endothelial cells Claudin-5 (CLDN5), CD31 (PECAM1) and vimentin (VIM) (**Supplemental Figure 1B**). Finally, we assessed the purity and morphology of the isolated cerebrovasculature through light microscopy. This examination unveiled sizable blood vessels that align with the characteristics of brain vasculature (**Supplemental Figure 1C**).

Using TMT-MS based quantitative proteomics we analyzed a total of 67 individual vessel preparations from postmortem frontal cortical tissues from the University of Pennsylvania School of Medicine Brain Bank (UPenn cohort) (**Figure 1A, Supplemental Table S1**). Importantly, in this study we also leveraged bulk proteomes from cortical samples from the same individuals profiled using an identical TMT-MS approach to better understand the relationship across bulk and vascular enriched proteomes (**Figure 1B**). A unified semi-quantitative method to assess both amyloid (CERAD) and tau (Braak stage) deposition was used to classify all cases, defined by these postmortem pathological and also clinical criteria, as neurologically healthy controls, AD, and PSP (UPenn cohort: *N* = 26 controls, 22 AD, and 19 PSP) (**Supplemental Table S1**). To characterize the vascular enriched fractions, we first evaluated contributions of core brain cell types to each proteome using a marker list extracted *in silico* from the data of a previously generated single-cell and single-nuclei RNA-seq study (**Supplemental Table S2**)^23^. This analysis showed that the vascular proteome was enriched in endothelial cells in contrast to other brain cell types, with the notable exception of five samples spanning various diagnostic groups. These outliers displayed minimal enrichment of endothelial cells but demonstrated a significant increase in neuron-specific proteins, and thus were excluded from subsequent analyses (**Supplemental Figure 2A**). Following outlier removal we adjusted for the confounding effects of age, sex, and postmortem interval (PMI)^29^. Moreover, in order to account for biases in sample fractionation, we normalized for neuronal-specific cell type signatures across all samples. This process resulted in the ultimate quantification of 9,854 vascular-enriched proteins across individual brain samples from controls, AD, and PSP groups (**Supplemental Figure 2B** and **C**). Comparably, the subsequent bulk proteome analysis led to the identification of 9,418 proteins. Among the identified unique protein gene products (rather than protein isoforms represented in the above counts) in both vascular and bulk proteomes, 1,183 were unique to the vascular proteome, 782 were only identified in the bulk proteome and the remaining 8,636 proteins showed overlap between both fractions (**Supplemental Figure 3A**). To investigate a relationship between both fractions in AD and PSP, we compared the effect size (log_2_ fold change) of quantified proteins in bulk and vascular proteomes across diagnosis groups, overlaying log_2_ fold enrichment in vasculature over bulk tissue protein (**Figure 1C** and **D, Supplemental Table S3** and **S4**). This revealed a partial overlap between proteomic profiles in AD relative to control, with an enrichment in both proteomes of amyloid-associated proteins, that are related to the matrisome such as COL25A1, MDK, NTN1, SFRP1, SMOC1, HGF, SPOCK3 and reduction of neuronal-specific proteins, including PCSK1, VGF, and NPTX2. Interestingly, proteins associated with SMCs and pericytes (DES, PDLIM3, VIM, PTN, and OLFML3) showed an opposite directionality of change with their higher abundance in the AD bulk proteome and concomitant decrease in AD vasculature (**Figure 1C**). Vascular and bulk proteomic signatures discovered in PSP showed higher consistency in directionality of proteins compared to AD. More specifically, we found an increase in SAA1, FGA, C4BPA, CA1, and HP proteins and a decrease in many fibroblast-specific proteins (COL1A1, COL1A2, LUM, OGN) (**Figure 1D**).

**Figure 1.**
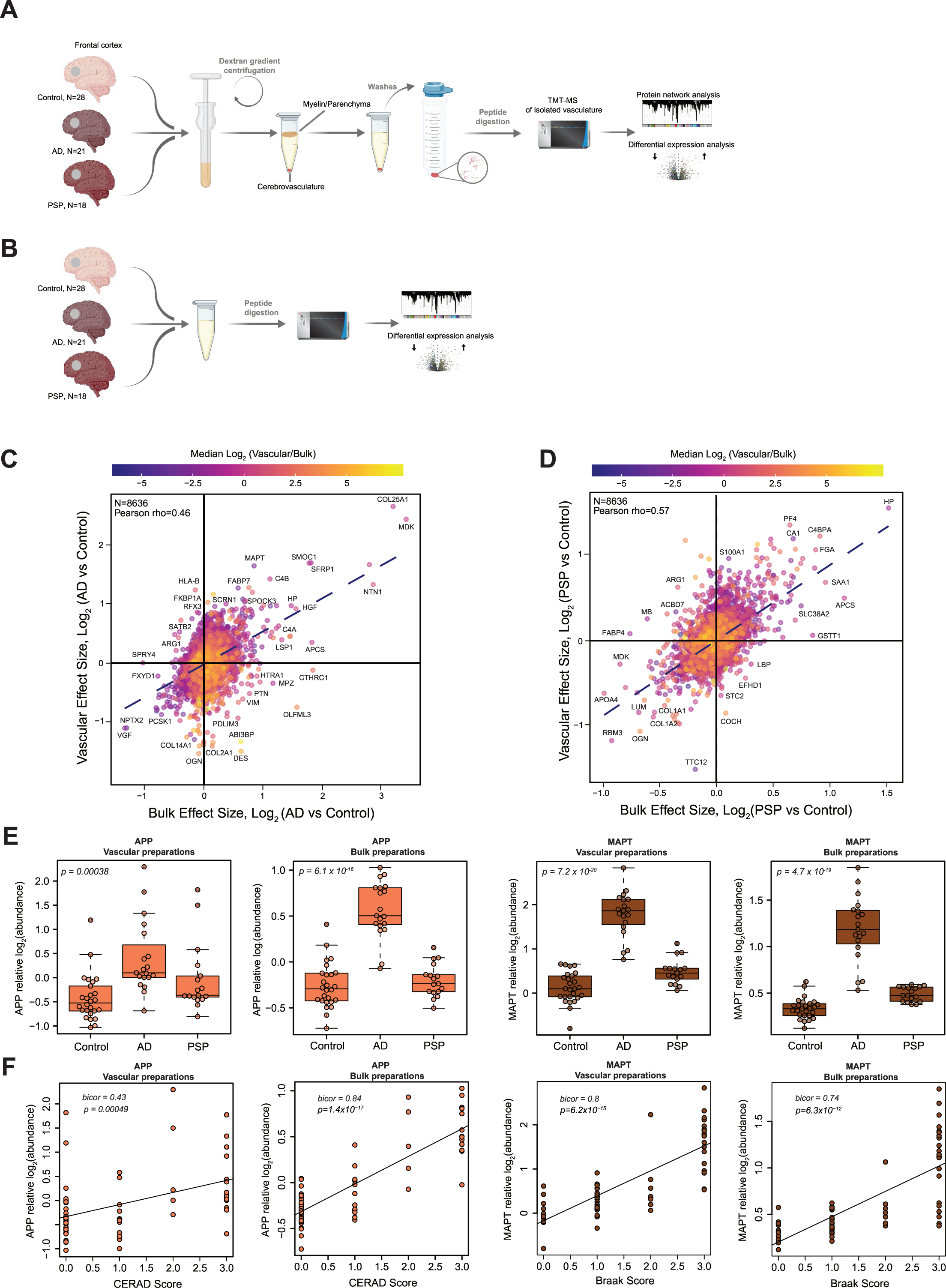
Deep quantitative proteomics of human cerebrovasculature and whole brain tissue. **(A-B)** Schematic representation of the experimental workflow for matched human postmortem tissues from non-demented controls (*N* = 28), AD (*N* = 21), and PSP (*N* = 18) cases from the University of Pennsylvania Brain Bank (UPenn) cohort. Isolated cerebrovasculature was obtained by dextran gradient centrifugation that allowed the separation of parenchyma/myelin from blood vessels. Both vascular and bulk fractions underwent enzymatic digestion followed by labeling with isobaric tandem mass tags and liquid chromatography mass spectrometry (TMT-MS). **(C-D)** Scatter plot of the log_2_ effect size in the bulk and vascular proteome in AD vs control **(C)** and PSP vs control **(D)**; color scale indicates the mean log_2_ (cerebrovasculature/bulk) enrichment. **(E)** TMT-MS quantified levels of APP and MAPT in vascular and bulk preparations. *P* values were calculated using one-way ANOVA for overall groupwise difference. Box plots represent median, 25^th^ and 75^th^ percentiles while box whiskers extend to non-outlier measurements up to 1.5 times each nearest interquartile range. **(F)** APP and MAPT abundance levels were positively correlated to CERAD and Braak scores, respectively using biweight midcorrelation (Bicor). Student’s test for correlation significance *p* values are provided for each correlation with *p* < 0.05.

Utilizing a pairwise differential expression approach, we sought to identify individual proteins that distinguish the vascular and bulk proteomic datasets and also differentially expressed in AD or PSP compared to control cases. In the vascular fraction of AD, we observed significant increases in i) several transport proteins (ATP7B, SLC4A11, APOF), ii) proteins associated with the ECM (ADMATSL4, PAPPA, COL16A1), and iii) transcription factors with pivotal roles in angiogenesis and vascular homeostasis (FLI1, POU3F, NFTAC2), were significantly increased. Conversely, proteins that showed significant increases or decreases in AD and uniquely identified in bulk fractions in AD were associated with neuronal signaling and axon biology (ORAI2, SYT6, SYT10, GLDN, KLK6, TPH2, SHISA6) (**Supplemental Figure 3B** and **C**). Pairwise analysis of protein changes in PSP in the bulk or vascular fractions revealed several overlapping proteins with AD, including APOF, MLKL, SLC4A11, B3GAT2 providing evidence for existence of shared mechanism possibly related to the pathological accumulation of tau (**Supplemental Figure 3B** and **D**).

Given that proteomic assessment of amyloid precursor protein (APP) and microtubule associated protein tau (MAPT) provide surrogate measurements of Aβ and tau, respectively^34^, we next evaluated their TMT-MS quantified levels in vascular and bulk preparations across all disease groups. As expected, the APP levels quantified by TMT-MS were significantly elevated in AD compared to controls and positively correlated with CERAD score, but there was no change observed in PSP (**Figure 1E** and **F**). Interestingly, MAPT levels showed significant increase in AD cases compared to controls and high correlation with Braak stages in vascular and bulk preparations potentially suggesting association of tau with the vasculature. Although we also observed an elevation of MAPT in the both bulk and vascular fractions, the effect was less dramatic in PSP than in AD compared to controls, highlighting the expected lower pathological burden of tau in frontal cortex of PSP cases^35^ (**Figure 1E** and **F**). Given that Aβ40 is recognized for its enrichment in brain vasculature^36^ and Aβ42 is considered a predominant form found in brain parenchyma^37^, we further examined the MS-quantified levels of the unique C-terminal tryptic peptides (Aβ42: GAIIGLMVGGVVIA and Aβ40: GAIIGLMVGGVV) from both these amyloid species in both the bulk and vascular fractions. Our findings indicate that the isolated cerebrovasculature exhibited an >4-fold enrichment of Aβ40, while >4-fold levels of Aβ42 were observed in the bulk fraction of the brain compared to the vascular fraction. Furthermore, the elevated levels (>2-fold) of the Aβ40/ Aβ42 ratio in AD cases compared to control and PSP within the vascular fractions align with previous evidence linking higher levels of Aβ40 to vascular CAA^38^ (**Supplemental Figure 4A**). In summary, AD demonstrated both shared and distinctive patterns of protein alterations in the vascular and bulk fractions with amyloid-associated proteins linked to the matrisome displaying the most significant effects. The overall magnitude of protein changes was notably lower in PSP compared to AD, aligning with the spatially distributed pathology observed in the frontal cortex in these cases.

### Cerebrovascular proteomic network analysis identifies modules associated with brain vascular cell types and functions

Our previous work has identified a reproducible network of highly preserved groups of correlated proteins termed modules from bulk tissue that established a biological context for the human AD brain proteome and allowed discovery of new pathophysiological processes related to disease^29,30^. To identify such biologically meaningful co-expression patterns in the cerebrovascular proteome, we applied an analogous network-based approach to the vascular-enriched proteome (**Figure 2A**). The constructed network highlighted expression of 93 protein modules across control and disease brain tissues, ranging in size from the largest module, M1 with n=812 protein members to the smallest module M93 with n=33 members (**Supplemental Table S5**). Gene ontology (GO) analysis was used to assign principal biology and specific molecular functions to each protein module using top GO terms (**Figure 2A, Supplemental Table S6**). Given that cellular composition can influence protein co-expression^28^, we assessed the cell type nature of each protein module using cell type specific protein markers derived from scRNA-seq studies for enrichment of endothelial cells, pericytes, SMCs, fibroblasts, astrocytes, microglia/macrophage, neurons, oligodendrocytes, and oligodendrocyte progenitor cells (OPCs) (**Supplemental Table S2**)^23^. In the present network we observed a significant relationship between module biology, disease status, and cell type specificity. Specifically, we found nine modules with prominent contribution of vascular cells, including three modules enriched with endothelial and pericyte markers: M16 ‘Cell Signaling’, M12 ‘Blood vessel development’, M5 ‘Maintenance of BBB’; four modules had abundant expression of SMC proteins, including M77 ‘Laminin/Endothelial cell (EC) proliferation’, M88 ‘MHC complex’, M28 and M90 Actin cytoskeleton/SMC contraction, and there were two modules that were associated with fibroblasts: M11 ‘ECM/Collagen fibril organization’, and M2 ‘Cell adhesion’ (**Figure 2A**). Using the ratio of vascular to bulk proteome to annotate a mean value for each vascular-enriched proteomic module, we next found an additional 25 modules comprised of proteins substantially enriched in the vasculature despite no obvious contribution of vascular cell types (**Figure 2A**). The analysis of paired vascular and bulk samples also allowed us to identify modules with novel proteins showing the strongest correlation to each module eigenprotein (i.e., ‘hub’ proteins). Importantly, we observed that modules enriched in cerebrovasculature had hub proteins that were previously unidentified or relatively depleted in the bulk proteome. This included M11 ‘ECM/Collagen fibril organization’, M35 ‘ECM organization’, M14 ‘Keratinization’, M88 ‘MHC complex’, M28, M90 ‘Actin cytoskeleton/SMC contraction’, M5 ‘Maintenance of BBB’, M77 ‘Laminin/Endothelial cell (EC) proliferation’, and M2 ‘Cell adhesion’ (**Figure 2B**). Thus, characterization of the protein systems biology of isolated cerebrovasculature significantly diverges from the bulk analysis, leading to identification of novel vascular-specific protein modules highlighting biological heterogeneity.

**Figure 2.**
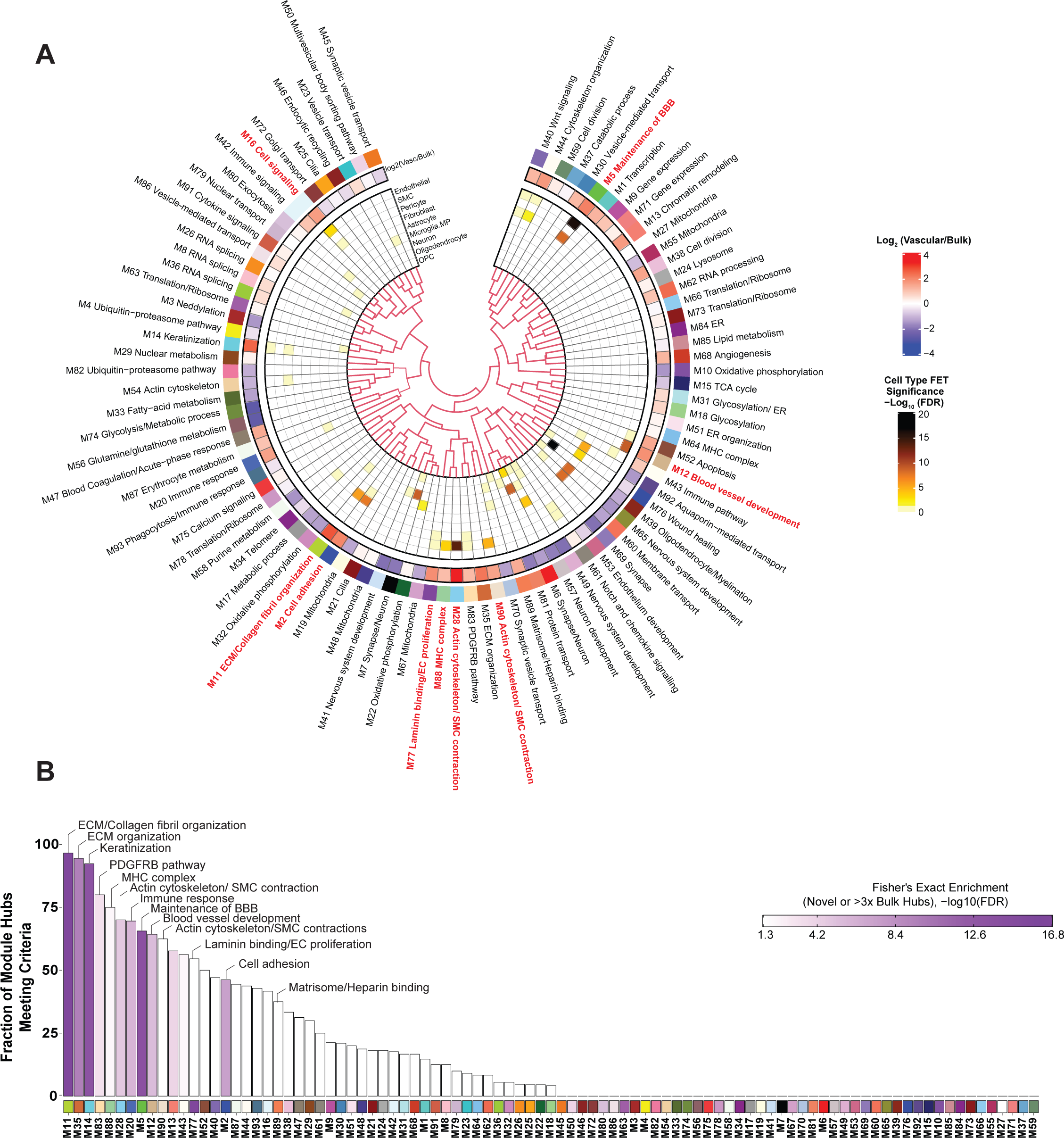
Protein co-expression network reveals novel modules enriched in cerebrovasculature. **(A)** Protein co-expression network of cerebrovasculature was built using WGCNA as described in Methods, and consisted of 93 protein co-expression modules (N = 9,854 proteins, each module represented by a different color and number for decreasing rank size). Gene ontology (GO) analysis was used to identify representative the modular biology (outer track labels). Mean log_2_ ratio of vascular to bulk proteomic abundance was performed to identify modules enriched in vascular proteome. The cell type association of each module was assessed by hypergeometric overlap of each module’s gene products with the cell type specific marker list for each brain cell type de novo extracted from single-nuclei RNA-seq data^23^, curated into 9 primary cell types; one-sided Fisher’s exact test with Benjamini-Hochberg correction was employed, with colors other than white representing significant FDR < 0.05 (inner panel). Scale bars indicate log_2_ ratio of vascular to bulk proteome (red indicates enrichment and blue indicates depletion) and cell type enrichment (from modestly significant to darker brown indicating stronger enrichment). Module relatedness is shown in the central dendrogram. (**B**) Top percent novel hubs identified in the TMT cerebrovascular network novel or ≥3-fold enriched relative to paired bulk protein measurements on average. Color scale represents gene product enrichment calculation using Fisher exact test with Benjamini-Hochberg FDR correction.

To further assess whether the protein modules identified in the vascular proteome were also present in the bulk protein network, we performed network preservation statistics, where the Z_Summary_ composite statistic indicates the degree of preservation of a module across networks. We found that 74 protein modules in the vascular network were preserved in bulk proteome with 17 modules showing high preservation (Z_Summary_ >10 or *p*<1×10^-23^) and 57 modules showing moderate preservation (Z_Summary_ >1.96 or *p*<0.05) indicating the robustness of the majority of the modules across both proteomes derived from fractionation or the same bulk tissue (**Figure 3**). Consistent with previous findings^29^, among the highly preserved modules we found protein communities commonly associated with AD pathophysiology, including M6 and M7 ‘Synapse/Neuron’, M39 ‘Oligodendrocyte/Myelination’, M10, M32 ‘Oxidative phosphorylation’, and finally M55 ‘Mitochondria’. Importantly, we also observed that 19 modules were only present in the vascular and not in the bulk network. Of these, 10 modules were enriched in cerebrovasculature, including M86 ‘Vesicle-mediated transport’, M14 ‘Keratinization’, M2 ‘Cell adhesion’, M77 ‘Laminin binding/EC proliferation’, M88 MHC complex, M90 ‘Actin cytoskeleton/SMC contraction’, M12 ‘Blood vessel development’, M18 ‘Glycosylation’, M31 ‘Glycosylation/ER’, M30 ‘Vesicle-mediated transport’, and M40 ‘Wnt signaling’ (**Figure 3**). This suggests that bulk proteomic analyses do not entirely capture differences on a network level in brain vasculature, emphasizing the significance of employing enrichment approaches to capture vascular changes in the brain proteome.

**Figure 3.**
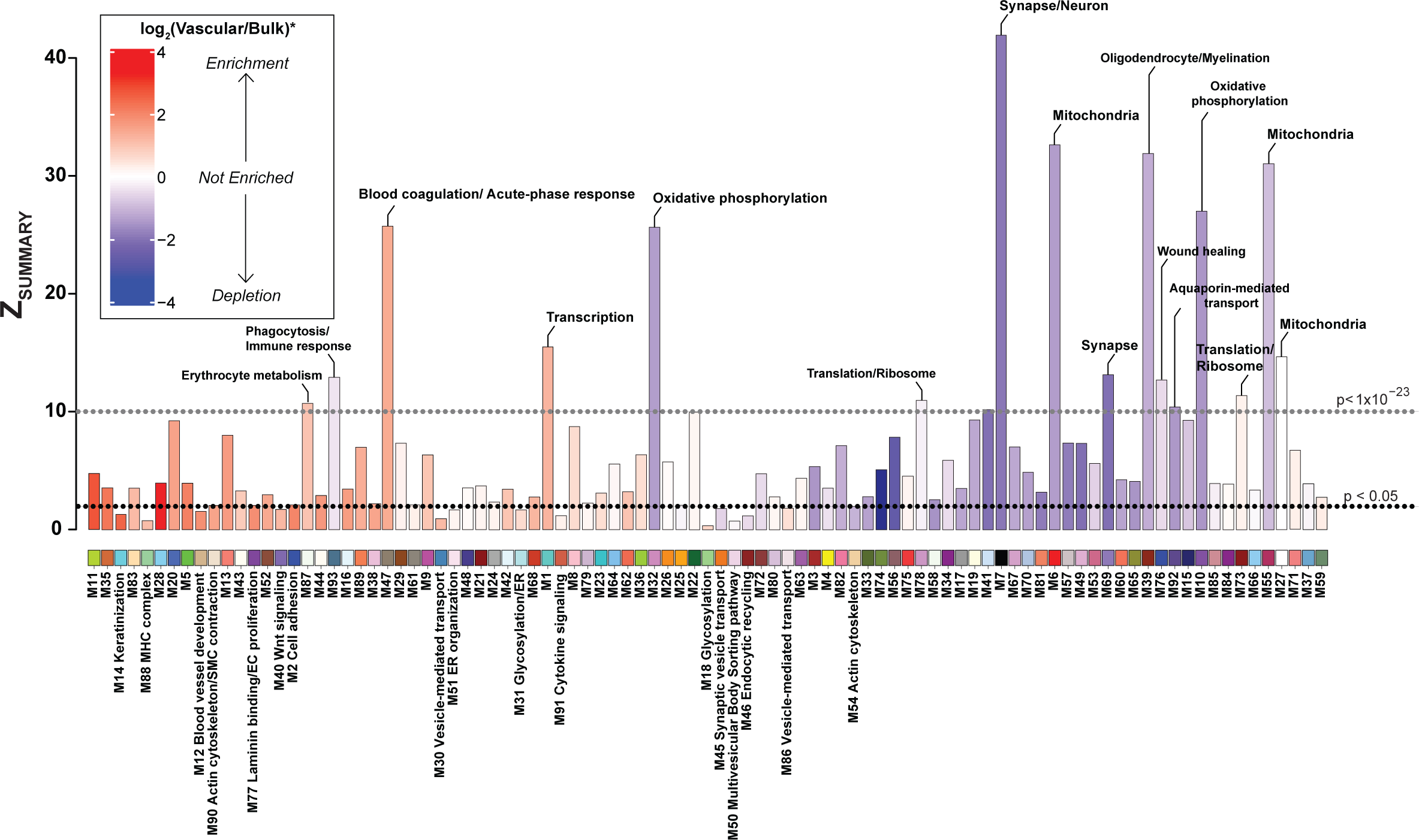
Cerebrovascular network preservation. Module preservation of the TMT protein vascular network into the proteomes of paired bulk tissue case samples from frontal cortex of matched individuals. Modules with a preservation Z_summary_ score less than 1.96 (*p* value<0.05) were not considered preserved. Protein modules with Z_summary_ score above 1.96 were considered moderately preserved (modules above black line), whereas protein modules with Z_summary_ score above 10 (*p* value< 1×10^-23^) were considered highly preserved (above grey line). Each bar is shaded according to the scale of mean pair cerebrovascular enrichment. Modules that showed no preservation in the bulk proteome but were enriched in vasculature are labeled in red.

### Correlation of cerebrovascular proteomic modules with distinct neuropathological phenotypes in AD and PSP

Given the robustness of our cerebrovascular network, we set out to uncover pathophysiological alterations associated with protein modules in AD and PSP. To this end, we first correlated modules with disease state and neuropathological hallmarks, including amyloid and tau burden presented as CERAD and Braak scores, respectively, as well as CAA severity score, gliosis, and *APOE4* genotype. We found that 66% of protein groups were strongly associated with AD and/or PSP and correlated with at least one neuropathological trait, whereas 34% of modules showed no substantial change in the disease conditions compared to controls. Specifically, among significantly changed modules 19 were altered only in AD, and 31 modules showed the same direction of change in AD and PSP, yet only 4 modules were different in PSP compared to controls, but not in AD (**Figure 4A, Supplemental Table S7**).

**Figure 4.**
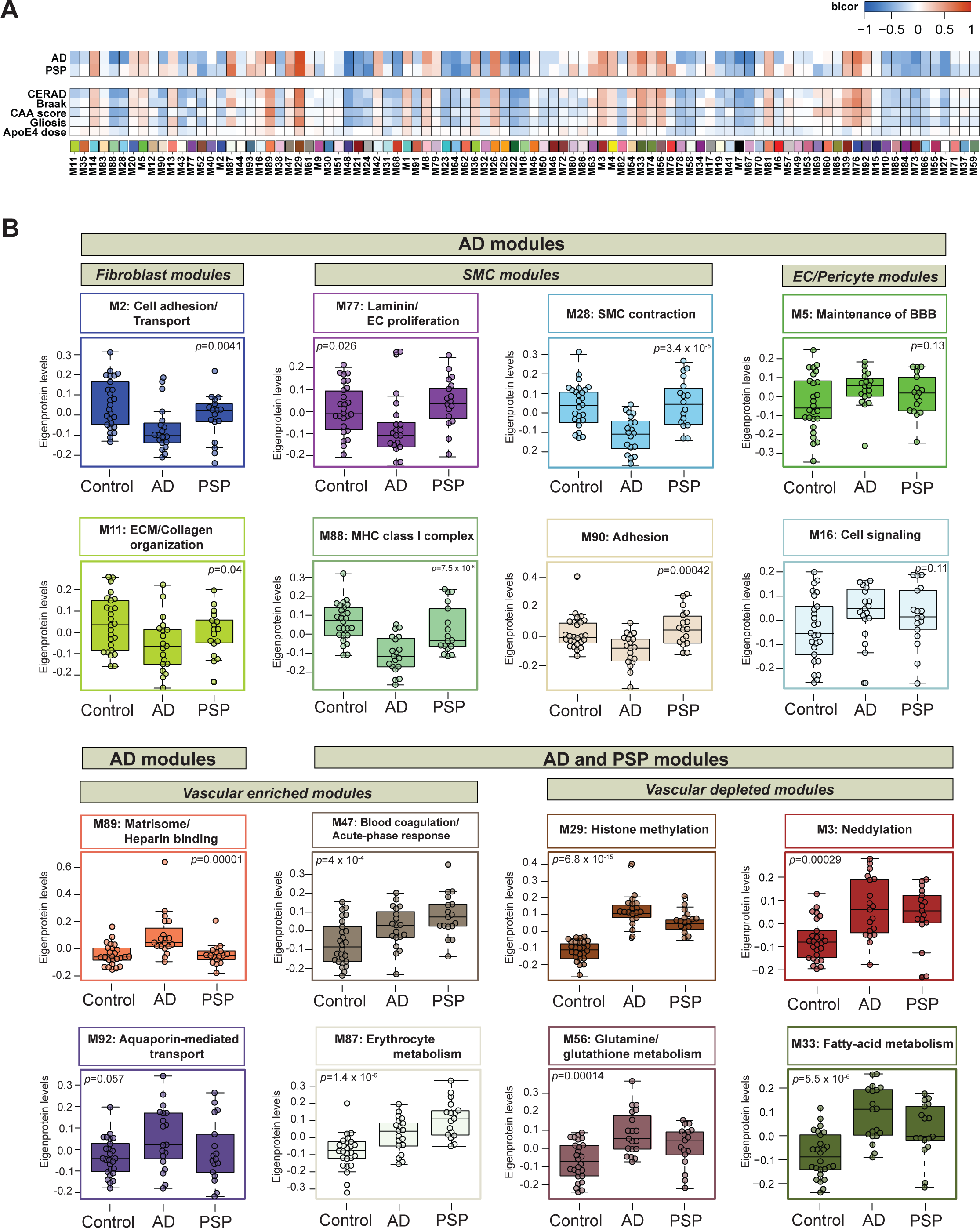
Co-expression network of cerebrovascular proteome resolves different proteomic signatures in AD and PSP. (**A**) The WGCNA co-expression network constructed from 9,854 proteins yielded 93 co-expression protein modules. The heatmap shows the correlations of module eigenproteins with binary disease/control status, neuropathological hallmarks (CERAD, Braak, CAA, gliosis), and *APOE Ɛ4* dose. Two-color heatmap represents strength of positive (red) or negative (blue) correlation, with *P* values provided for all correlations with *P* < 0.05. (**B**) Module eigenprotein (ME) levels grouped by case status were plotted for protein modules of interest. MEs were analyzed using one-way ANOVA test and unadjusted *p* values are provided for each module. Box plots represent median, 25^th^ and 75^th^ percentile while box whiskers encompass actual data points up to 1.5 times the nearest interquartile range.

Next, we assessed differences between disease states across the various vascular-relevant biological systems. The evaluation of eigenproteins, which reflect the first principal component expression profile for all proteins of a module, yielded two main clusters based on their association with disease class and vascular contribution. The first group exhibited significant changes in AD frontal cortex, but not in PSP when compared to controls (**Figure 4A** and **B**). It included modules strongly associated with vascular cells. Among these, six modules, including M11 ECM/Collagen fibril organization, M2 ‘Cell adhesion’ M77 ‘Laminin/Endothelial cell proliferation’, M88 ‘MHC complex’, M28 and M90 ‘Actin cytoskeleton/SMC contraction’ demonstrated a significant enrichment with fibroblasts and SMCs and showed negative correlation with neuropathological traits, particularly CERAD and CAA score, reflecting the specificity of vascular changes in response to amyloid pathology in AD. Modules M5 ‘Maintenance of BBB’ and M16 ‘Cell signaling’ were largely enriched with endothelial and pericyte functions, but displayed only a modest increase in the abundance levels in AD, possibly due to capturing less prominent alterations affecting capillaries compared to larger vessels in our cohort. This group also featured a module linked to ‘Matrisome/Heparin binding’ (M89) that showed a significant elevation in AD (*p* = 0.00001) and positive correlation to amyloid and tau deposition in the brain.

The second group comprised of biologically diverse modules with a significant increase in both AD and PSP and was further divided into modules depleted in blood vessels such as M3 ‘Neddylation’, M29 ‘Nuclear metabolism’, M33 ‘Fatty-acid metabolism’, and M56 ‘Glutamine/glutathione metabolism’ or enriched in the cerebrovasculature, including M47 ‘Blood coagulation/Acute-phase response’ and M87 ‘Erythrocyte metabolism’. Of note, M47 and M87 were the only modules in this group that displayed strong association with Braak scores but no correlation to amyloid burden suggesting the interplay between tau pathology and these vascular-related functional changes (**Figure 4A** and **B**). Collectively, cerebrovascular proteome network analysis revealed novel, heterogeneous biology influenced by hallmark pathological features of AD and PSP, and in many cases linked to brain vascular cells.

To examine molecular signatures of isolated cerebrovasculature distinguishing AD and PSP, we set out to identify significant protein alterations in each disease group compared to non-demented controls (**Figure 5A** and **B**). AD exhibited a larger number of differentially altered proteins than PSP in relation to controls, possibly due to more prominent structural and functional changes that occur due to amyloid deposition in cerebrovasculature in AD frontal cortex^39^. Pairwise comparison of AD to controls outlined a large fold increase in M89 ‘Matrisome/ Heparin-binding’ proteins, such as SMOC1, MDK, C4B, SFRP1, and NTN1 with high specificity even as these proteins were not differentially expressed in the PSP versus control comparison. This protein signature, previously mapped to a bulk Brodmann area 9 “Matrisome” module^29^, has been shown to consistently distinguish AD from other proteinopathies. These proteins either directly bind to amyloid or are associated with amyloid plaques, highlighting their crucial connection to amyloid deposition in the brain^29,33^. As expected, MAPT was not uniquely altered in AD but showed higher elevation in AD than PSP, reflecting higher burden of tau pathology in frontal cortex in AD cases. The remaining signature of increased proteins in AD was associated with nuclear metabolism (SDC4, SPOCK3, CEP63) and fatty-acid metabolism (FABP7, TP22, and FKBP1A) (**Figure 5A**). Although, our previous study identified SDC4 and SPOCK3 as members of the ‘Matrisome’ module in bulk fractions^29^, here these proteins were mapped to a module featuring MAPT, potentially suggesting their closer relationship to tau than amyloid in vasculature. In contrast, among proteins showing a steep reduction in abundance in AD, a number were involved in SMC function (ACTA2, TAGLN, TPM1, TPM2, PGM5) and extracellular matrix (DES, OGN, CPXM1, PAPPA, FMOD) which may suggest cell loss or subtype shifting in response to amyloid deposition in brain parenchyma and cerebral blood vessels (**Figure 5A**). In addition, consistent with previous reports^29^, neuronal markers with known links to synaptic plasticity, such as VGF and NPTX2 showed declining trends in AD. Markers significantly altered in PSP across pairwise comparisons prominently featured members of M47 ‘Blood coagulation/ Acute-phase response’, including SPTA1, SPTB, RHCE, APOB and M87 ‘Erythrocyte metabolism’, including APOF, C4BPA, C4BPB, and PROS1. In summary, the frontal cortex vascular-enriched network outlines key differences in protein signatures of AD- and PSP-associated pathologies enabling differentiation between different disease states (**Figure 5, Supplemental Figure 5**).

**Figure 5.**
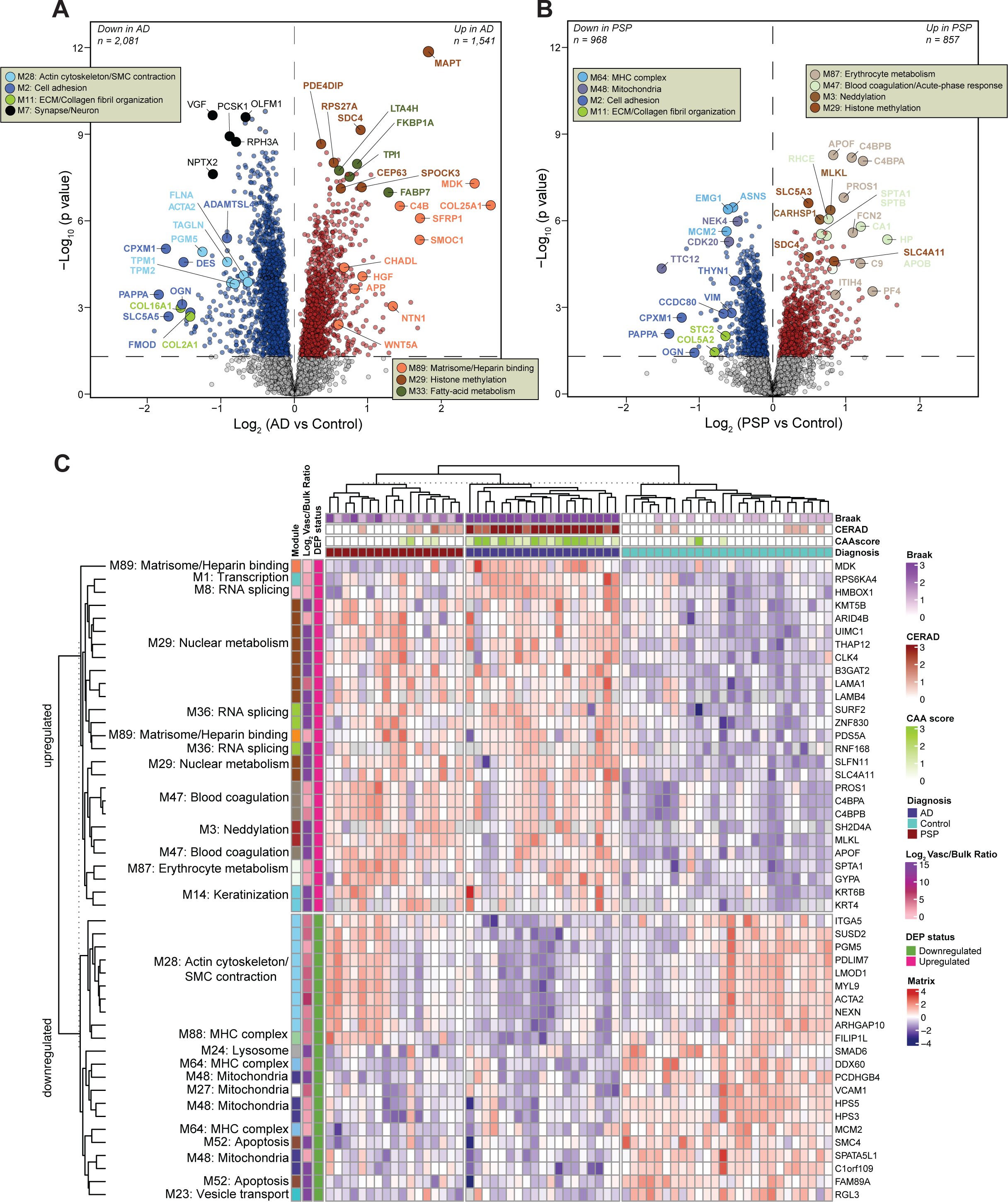
Disease status demonstrates different cerebrovascular protein signature. (**A**-**B**) Volcano plots showing differential abundance of proteins between control and AD groups (*N* = 3,622) (**A**) or proteins between control and PSP cases (*N* = 1,825) (**B**). The x axis illustrates the log_2_ fold change (AD vs Control, **A**) or (PSP vs Control, **B**), while the y axis represents the -log_10_ statistical *p* value calculated for all proteins in each pairwise group. *P* values were obtained from one-way ANOVA with Tukey’s post hoc test and for Tukey *p* values less than 10^-8.5^, Bonferroni correction of two-tailed unequal variance t-test *p* values replaced highly significant but imprecisely estimated Tukey *p* values. Proteins significantly increased in AD (*N* = 1,541) (**A**) or PSP (*N* = 857) (**B**) are depicted in red (*p* < 0.05) whereas those significantly decreased in AD (*N* = 2,081) (**A**) or PSP (*N* = 968) (**B**) are highlighted in blue. Proteins of interest are shown as enlarged dots and shaded according to color of the module membership. Grey shaded dots represent proteins with unchanged levels. (**C**) Supervised hierarchical clustering of the 49 most significant proteins that were unique to vasculature or 3-fold enriched in vasculature compared to bulk and with volcano significance *p* < 0.05 across each of the three possible comparisons among control, AD, and PSP groups. Red shaded boxes highlight proteins upregulated and blue shaded boxes indicate downregulated proteins relative to the central tendency. Minimum and maximum log_2_ (abundance/central tendency) are clipped at – and +4.

To determine if these differentially expressed proteins could function as classifiers for disease status and pathological abnormalities, we conducted a supervised clustering approach. This analysis was confined to the 49 most significant proteins that were either unique to vasculature or at least 3-fold enriched in vasculature compared to bulk and also differentially expressed (ANOVA+Tukey *p*<0.05) across all three comparisons between control, AD, and PSP groups. Notably, AD was easily distinguished from controls by significant elevation of proteins associated with M89 ‘Matrisome/Heparin binding’, M47 ‘Blood coagulation/ Acute-phase response’, M87 ‘Erythrocyte metabolism’, M8 and M36 ‘RNA splicing’, M29 ‘Nuclear metabolism’, M3 ‘Neddylation’, and M33 ‘Fatty-acid metabolism’. AD also displayed significant decrease of proteins in modules largely linked to SMC biology, mitochondria, and metabolic processes compared to controls. (**Figure 5C**). PSP vascular-enriched samples showed a similar pattern to AD for increased proteins but a more heterogenous proteomic profile of decreased proteins compared to AD or controls (**Figure 5C**). Collectively, the selected differentially altered proteins provide an AD-specific signature that diverges from controls and PSP.

### AD GWAS candidate genes are overrepresented in modules enriched in cerebrovasculature

Integrated analysis of genetic association signals derived from large-scale genome wide association studies (GWAS) with the co-expression modules has a great potential to nominate pathways implicated in AD pathophysiology^28^. To assess whether proteins encoded by GWAS AD risk genes were enriched in any particular modules in our network, we analyzed the single nucleotide polymorphism (SNP) summary statistics for three AD GWAS (**Supplemental Table S8**)^40-42^ to calculate gene level association using three runs of the algorithm Multi-marker Analysis of GenoMic Annotation (MAGMA) as well as a PSP GWAS (**Supplemental Table S9**)^43^. The gene-level MAGMA summary *p*-values for disease risk were considered as an ensemble for all three AD GWAS. Our vascular network featured five modules with overrepresentation of proteins from AD GWAS candidate loci, including M11 ‘ECM/Collagen fibril organization’, M67 ‘Mitochondria’, M77 ‘Laminin/Endothelial cell proliferation’, M89 ‘Matrisome/Heparin binding’, and M5 ‘Maintenance of BBB’ (**Figure 6A**). Strikingly, M11, M77, M89, and M5 modules showed significant enrichment in human cerebrovasculature, further underscoring the importance of vascular dysfunction in pathogenesis of AD. While a total of 34 candidate risk genes mapped to M11 ‘ECM/Collagen fibril organization’, including *APOC1*, *APOC2*, *PVR*, and *GEMIN1*, contribution of 14 gene products led to enrichment of M77 ‘Laminin/Endothelial cell proliferation’, including BCAM, CAV1, and CAV2. The strong association of M89 with AD was largely driven by the presence of *APOE*, the remaining 14 risk-associated genes showing less contribution to the module enrichment, including *KNOP1*, *APP*, *CHADL*, *SMOC1*, and *SMOC2*. Finally, M5 ‘Maintenance of the BBB’ constituted of 83 candidate risk-associated genes with mostly small effect size, with the exception of *NECTIN1* that showed an overweighted significance of contribution to the enriched risk in the module.

**Figure 6.**
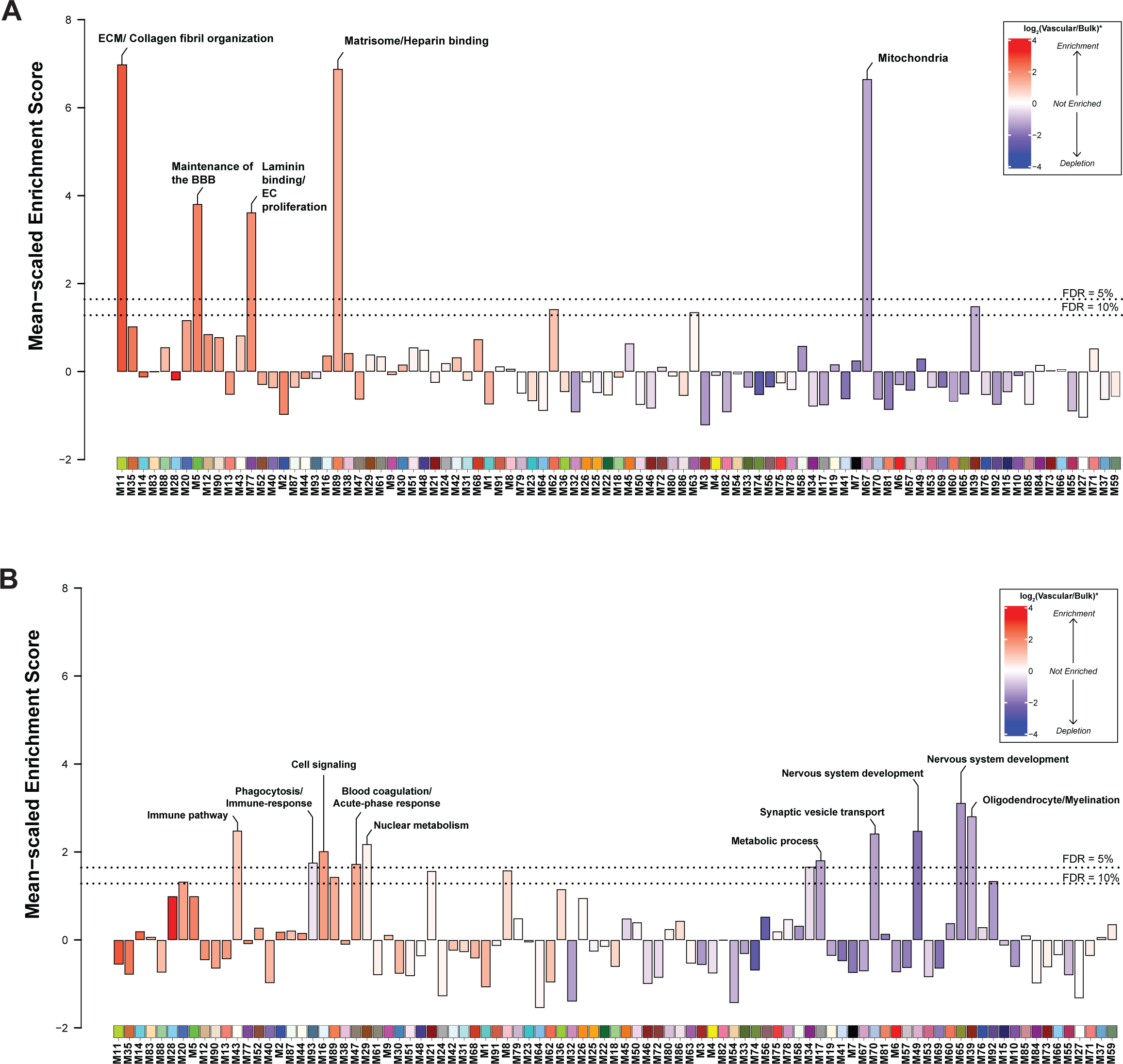
AD GWAS risk genes are overrepresented in protein modules enriched for brain vascular cells. **(A)** AD GWAS candidate genes were significantly enriched in five modules, including M11, M67, M77, M89, and M5. These modules were significantly up- or downregulated in AD, as shown in Figure 2A. The horizontal dotted lines indicate the thresholds for permutation test false discovery rate (FDRs of 10 or 5 percent), above which genetic risk enrichment was considered significant. Bar colors indicate mean all-protein module member mean log_2_(cerebrovasculature/bulk) proteome fractionation enrichment. **(B)** GWAS risk genes for PSP were mapped to 10 modules with 5% FDR, among which two were significantly upregulated in PSP, including M29, and M47 (Figure 2B).

Gene-level risk enrichment of PSP causally associated gene products by module also identified candidate genes with MAGMA *p*-values< 0.05, and whose net risk was also significantly overrepresented in 10 modules. As expected, modules enriched for PSP GWAS risk genes differed from AD and were comprised of 4 cerebrovascular-specific modules, including M16 ‘Cell signaling’, M29 ‘Nuclear metabolism, M47 ‘Blood coagulation/ Acute-phase response’, and M43 ‘Immune pathway’ (**Figure 6B**). We did not find single candidate risk genes with overweighted significance driving the PSP GWAS risk association with protein modules. Instead, we observed modest contributions from multiple risk-associated genes, each contributing equally to the overall module enrichment of risk. Specifically, M16 featured 33 candidate genes, including *CYTH1, AFAP1L2,* and *MICALL1*, and M43 consisted of 30 gene products such as ACOX1, PLOD3, and IKBIP). Furthermore, M29 exhibited overrepresentation of genetic risk with 30 candidate risk-associated gene products, like B3GAT2, ARID4B, SDC4, and COL15A1, and finally M47 harbored 37 nominally significant PSP risk genes, including *PF4, FBXW7, SERAINEF2*, and *A2M*. Notably, we previously observed a significant increase in the abundance of M29 and M47 eigenproteins in PSP, further indicating the importance of these modules changing in disease development or progression. The other 6 modules enriched with PSP risk gene product proteins were depleted in cerebrovasculature but displayed significant enrichment for PSP GWAS were associated with neuronal and synaptic function, myelination, and metabolic processes (**Figure 6B**). In summary, these findings offer network-based evidence and organization of potential genetically linked factors contributing to vascular-associated changes occurring in AD or PSP, underscoring the critical role of cerebrovasculature in the etiology and pathophysiology of these diseases.

### Matrisome proteins are present in cerebral blood vessels and associate with CAA

As we consistently identified matrisome/heparin-binding proteins as highly specific and sensitive discriminators of AD in both bulk and vascular proteomes, our focus turned to understanding their spatial relationship with neuropathological hallmarks of AD. Module M89, associated with the matrisome, was comprised of 39 members (**Figure 7A**), of which the vast majority of proteins were mapped to this module for the first time. Only 10 proteins of M89 overlapped with the previously identified Matrisome module (M42) in a consensus bulk proteome of AD brain^29^, including SMOC1, NTN1, MDK, SFRP1, SLIT2, APP, APOE, NXPH1, TMEFF2, and SLIT1. In contrast, in depth quantification of cerebrovasculature enabled us to identify additional proteins linked to angiogenesis, ECM and SMC functions that overlap with ECM organization and collagen fibrillization. These M89 proteins included HGF, SLIT3, CHADL, ANOS1, SMOC2, HHIPL1, B4GALNT1, and CCN1, among others. As expected, the products of the APP processing, including Abeta40 and Abeta42 were also found in M89. Furthermore, we found that a few members of the previously described M42 ‘Matrisome’ module in bulk tissue, including SDC4, COL25A1, and SPOCK3, mapped to M29 (‘Nuclear metabolism’), which showed a significant correlation with both AD and PSP in our network. This suggests an association of M29 with tau-related changes. Other matrisome module proteins in bulk tissue, such as CTHRC1, OLFML3, FRZB, and COL11A1, were assigned to M11 in our proteome. Despite its association with ECM and collagen fibrilization, M11 exhibited a decrease in AD isolated vasculature, in contrast to the increase observed in M89.

**Figure 7.**
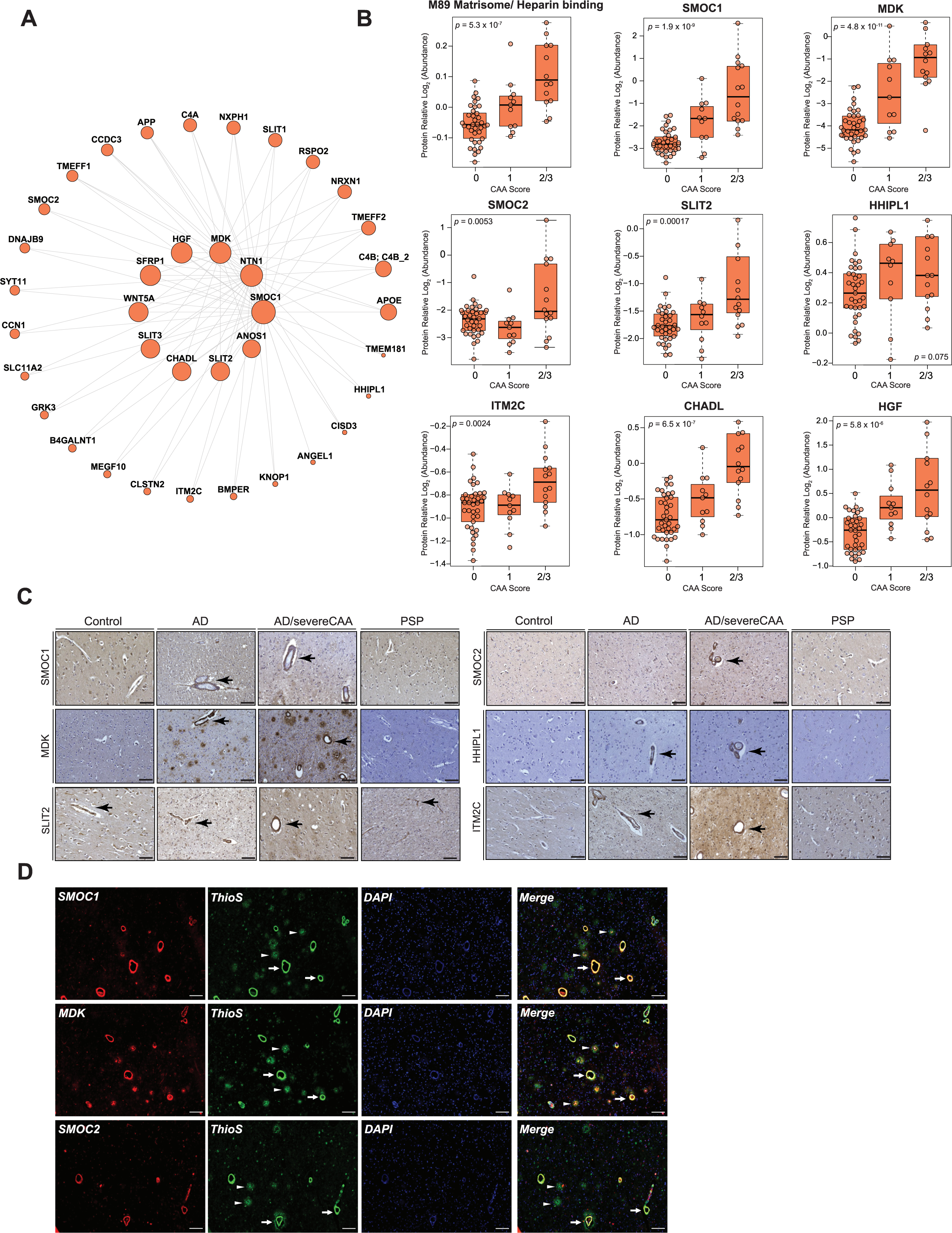
Matrisome proteins are found in cerebrovasculature in AD and associated with CAA. **(A)** An iGraph represents the top 50 proteins identified in the M89 Matrisome/Heparin binding module. Lines between proteins represent topological overlap matrix weight corresponding to the similarity of correlated patterns of node pairs over the 62 case samples in the cerebrovascular network. **(B)** Protein abundance of module M89 and selected module members across different CAA severity score. A 0 indicates no CAA, 1 indicates mild CAA, and 2/3 indicates moderate to severe CAA score. One-way ANOVA *p* values were assessed for each 3-group box plot. Box plots represent the median and 25th and 75th percentiles, with data points up to 1.5 times the nearest interquartile range beyond the box defining the extent of error bar whiskers. **(C)** Immunohistological evaluation of expression patterns of SMOC1, MDK, SMOC2, SLIT2, HHIPL1, and ITM2C in human postmortem cortical tissues from non-demented control, AD, AD with severe CAA, and PSP. Arrows indicate the presence of these proteins in the cerebrovasculature, the arrowheads indicate their expression in brain parenchyma. Scale bar, 100 μm. **(D)** Co-localization of SMOC1, MDK, and SMOC2 with amyloid in brain parenchyma and cerebrovasculature in human AD brain tissue. Thioflavin-S (thioS) stain was used to label fibrillar amyloid deposition. Arrows indicate the co-localization of these proteins with CAA, the arrowheads indicate their co-localization with amyloid plaques. Scale bar, 100 μm.

We next assessed whether previously and newly discovered module-member proteins were associated with CAA by plotting the selected M89 markers to each case sample’s CAA burden score, including no CAA (0), mild CAA (1), and moderate to severe CAA reflected by CAA scores of 2 and 3 (**Figure 7B**). We found that SMOC1, MDK, CHADL, and HGF displayed a stepwise increase in their abundance in relation to the severity of CAA. Meanwhile, SMOC2, SLIT2, and ITM2C showed a significant elevation in cases only presenting with moderate to severe CAA (**Figure 7B**). To further corroborate these findings, we employed immunohistochemistry to reveal the spatial distribution patterns in the frontal cortex across different disease states. Notably, our analysis revealed abundant accumulation of SMOC1, MDK, SLIT2, SMOC2, HHIPL1, and ITM2C localized to cerebral blood vessels in AD and AD presenting with severe CAA. This suggests that these proteins co-deposit with amyloid in the walls of cerebral vessels. Intriguingly, SLIT2 was also detected in cerebrovasculature in controls and PSP, indicating a specific basal vascular distribution pattern for this protein even in cases lacking vascular pathologies (**Figure 7C**). This was further supported by strong association of SMOC1, MDK, and SMOC2 expression with amyloid deposition in vasculature as CAA, indicated by thioflavine-S stain (**Figure 7D**). Although, SMOC1 and MDK also co-localized with fibrillar amyloid plaques in brain parenchyma, SMOC2 was nearly exclusively present in the Aβ-laden vessels, indicating its preferential vascular distribution pattern (**Figure 7D**). In summary, these findings highlight the discovery of new amyloid-associated proteins in the cerebrovasculature co-expression module, termed here ‘Matrisome/Heparin binding’ M89 module, which were not previously mapped to a ‘Matrisome’ module in bulk fractions^29^. Importantly, the heightened signature of module members aligned with the prominent co-occurrence of CAA, demonstrates a high level of specificity of this novel proteomic signature in amyloid-related neuropathological changes.

### Integrative analyses of brain and biofluid proteomes reveal biomarkers reflective of cerebrovascular changes in AD

Given that cerebrovascular dysfunction contributes to AD pathogenesis, and more than 90% of individuals with AD present with concurrent cerebrovascular disease^39^, there is a strong rationale to identify peripheral biomarkers that reflect this underlying pathophysiological process. In this study, we employed an integrated analysis to identify plasma and CSF biomarkers that potentially reflect vascular changes in the brain. To this end, we leveraged previous datasets^44,45^ (**Supplemental Table S10** and **S11)** to map differentially abundant proteins in both AD plasma and CSF to the cerebrovascular network to prioritize peripheral protein signatures that were enriched in modules associated with vascular biology in brain (**Figure 8A** and **B**). Importantly, AD patients in these biofluid studies were confirmed to be biomarker positive by CSF ratios of Aβ42 and tau. Collectively, this includes two separate sets of plasma samples: set 1 (N = 36) and set 2 (N = 85) encompassing a total of 47 control and 62 AD samples in the plasma proteomics study (**Supplemental Table S10)**^44^ and a single CSF set comprising of 141 control and 140 AD samples in the CSF TMT-MS proteomic study (**Supplemental Table S11)**. Using each of these datasets independently, we identified brain vascular modules that were enriched with proteins significantly increased or decreased in AD CSF and plasma (**Figure 8A** and **B**, **Supplemental Table S12**-**S15**). Notably, this revealed a highly concordant and significant increase in many members of M89, the ‘Matrisome/Heparin binding’ module, including SMOC1 and SMOC2, in the brain, plasma, and CSF of individuals with AD (**Figure 8C-F**). Furthermore, we observed a high correlation between these proteins and well-established core biomarkers, including Aβ42, total tau, and phosphorylated tau at position 181 (pT181)^46-48^ (**Figure 9**), making them promising biomarkers of vascular dysfunction in the AD brain. Interestingly, we also found that other M89 members such as MDK demonstrated increased trends in cerebrovasculature and plasma, but displayed a significant reduction in CSF suggesting that other regulatory mechanisms may play a role in their levels across biofluids. We also identified protein modules displaying divergent expression trends across brain, CSF, and plasma, including M11 ‘ECM/Collagen fibril organization’, M35 ‘ECM organization’, M47 ‘Blood coagulation/Acute-phase response’, M3 ‘Neddylation’, and M33 ‘Fatty-acid metabolism’, M74 ‘Glycolysis/Metabolic process, M56 ‘Glutamine/glutathione metabolism’, M58 ‘Purine metabolism’ (**Figure 8**). More specifically, the proteins associated with brain fibroblast biology and ECM within M11 and M2 were notably decreased in the cerebrovasculature and CSF in AD, yet there was a striking elevation in these proteins in AD plasma. Indeed, proteins such as BGN, CHST15, COL12A1, DCN, and COL14A1, among others exhibited the highest levels in AD plasma and demonstrated a significant correlation with conventional pathological traits (**Figures 8** and **9**). In summary, integrative cross-fluid proteomic characterization proves to be a promising tool for prioritizing peripheral biomarkers linked to CAA and cerebrovascular dysfunction in AD.

**Figure 8.**
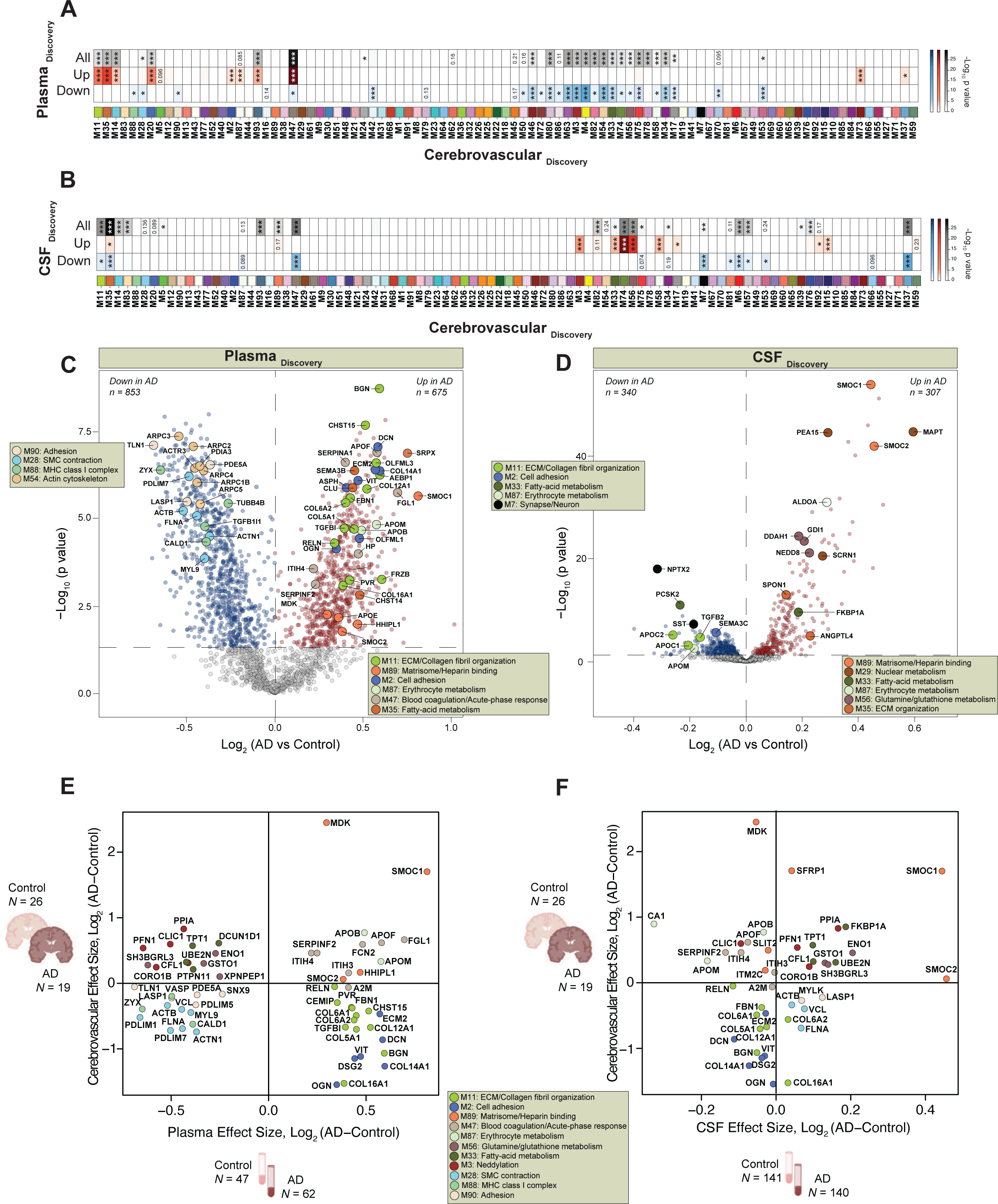
Integrated analysis of the brain vasculature and CSF and plasma proteomes in AD. (**A**-**B**) Overlap of proteins identified in the discovery cerebrovascular proteome (*N* = 26 control and *N* = 19 AD) with separate plasma cohort (*N* = 47 control and *N* = 62 AD) (**A**) or CSF (*N* = 141 control and *N* = 140 AD) (**B**) discovery datasets. In the plasma proteome *N* = 2,865 proteins were identified, whereas in the CSF proteome *N* = 2,067 proteins were quantified. Separate hypergeometric overlap Fisher exact analyses were used to assess the significance of protein overlap. The top row indicates overlap between brain vascular and plasma or brain vascular and CSF proteome (grey shaded scale). The middle row demonstrates the overlap between brain vascular and plasma or brain vascular and CSF proteins significantly increased in AD (*p*<0.05) (red shaded scale). The bottom row depicts the overlap between brain vascular and plasma or CSF proteins significantly decreased in AD (*p*<0.05) (blue shaded scale). The intensity of color shading indicates the degree of the overlap. Statistical significance is shown in the heatmap using stars (* FDR< 0.05, ** FDR < 0.01, and *** FDR < 0.001). FDR corrected values were determined using the Benjamini-Hochberg method. (**C**-**D**) Volcano plots showing differential abundance of proteins measured in plasma (*N* = 1,528) (**C**) or proteins measured in CSF (*N* = 647) (**D**) between Control and AD groups. The x axis illustrates the log_2_ fold change (AD vs Control), while the y axis represents the -log_10_ statistical *p* value calculated for all proteins in each pairwise group, obtained as Tukey post-hoc test p values following one-way ANOVA, except for imprecisely calculated Tukey values below 10^-8.5^ which underwent more precise and stringent Bonferroni post-hoc correction of a two-tailed unequal variance t-test. Proteins significantly increased in plasma in AD (*N* = 675) (**C**) or CSF in AD (*N* = 307) (**D**) are depicted in red (*p* < 0.05) whereas those significantly decreased in plasma in AD (*N* = 853) (**C**) or CSF in AD (*N* = 340) (**D**) are highlighted in blue. Proteins of interest are shown as enlarged dots and shaded according to the color of their module membership. (**E**-**F**) Scatter plots illustrate a Pearson correlation between log_2_ effect size (AD vs Control) of significantly altered proteins in cerebrovasculature and plasma (**E**) or cerebrovasculature and CSF (**F**). The significance of Pearson correlation was determined by Student’s *t*-test for significance of correlation implemented in the WGCNA R package.

**Figure 9.**
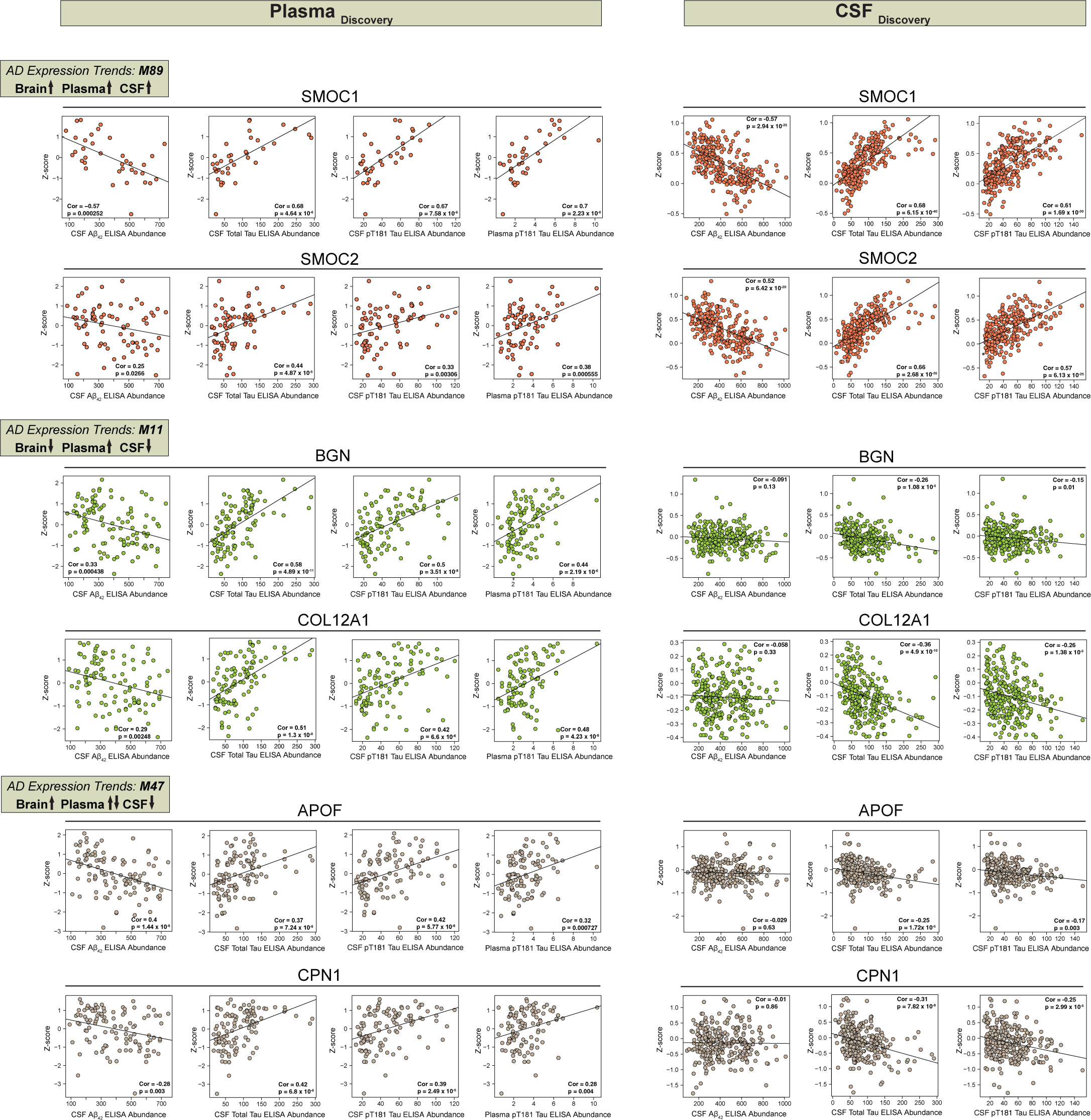
Specific cerebrovascular enriched proteins are detected in CSF and plasma, and strongly correlate with core AD biomarkers. Scatterplots represent the correlation measurements of CSF A(_42_, total tau and p-tau181 and plasma p-tau181 with six plasma (left panel) (total sample measurements: SMOC1 *N* = 36, SMOC2 *N* = 80, BGN, COL12A1, APOF, CPN1 *N* = 109) and CSF (right panel) proteins (total sample measurements for all six proteins *N* = 281). The Pearson correlation coefficient and Student’s *p* values are provided for each plot.

## Discussion

Detecting vascular changes in the brain within bulk tissues has proven challenging, limiting our ability to characterize proteomic alterations in less abundant cell types. In this study, we performed unbiased quantitative proteomics from distinct cortical fractions, including bulk and vessel-enriched samples obtained from the same individuals to identify pathways pertinent to cerebrovascular dysfunction in AD and/or PSP. Applying a co-expression network approach, we resolved 93 protein modules across controls, AD, and PSP individuals associated with heterogenous biological systems and cell types. Characterization of vascular and bulk proteomes revealed novel pathways relevant to vascular alterations in these neurodegenerative diseases related to amyloid and tau pathologies. In order to prioritize peripheral biomarkers for AD associated with vascular dysfunction in the brain, we assessed the overlap between proteins that show differential abundance in AD CSF and plasma, and network modules enriched in vascular-related proteins in brain; modules with mean protein enrichment in vascular preparations relative to their paired bulk samples were also identified and prioritized. This analysis highlighted the prominence of matrisome proteins, specifically SMOC1 and SMOC2, which exhibited increased levels in CSF, plasma, and brain tissues. Furthermore, immunohistochemical analysis revealed SMOC1 deposition in both parenchymal plaques and CAA in the AD brain, whereas SMOC2 was primarily localized to CAA. These findings collectively contribute significantly to our understanding of the involvement of cerebrovascular abnormalities in AD, providing insights into potential biomarkers and molecular pathways associated with CAA and vascular dysfunction in neurodegenerative diseases.

Recent advances in system-level proteomics have provided an invaluable view of the complexities of disease-relevant alterations in the human brain. Network-based analyses of AD bulk brain tissues demonstrated high reproducibility and consistency in identifying protein targets across various cohorts^29,30^. The depth of the TMT-MS pipeline facilitated the identification of modules linked to pathological endophenotypes in AD and PSP. These modules feature hub proteins not evident in corresponding bulk proteome analyses and were characterized by a vascular cell-specific signature. This implies the presence of distinct biological pathways governing cerebrovascular alterations in AD and PSP and underscores the significance of isolating the human vasculature for proteomic investigation.

The current dataset demonstrates the significance of employing targeted enrichment approaches when profiling less abundant brain cell types. Recent studies have adopted a similar approach to characterize the proteomic signature of isolated microvasculature in AD and CAA type 1^32^. Intriguingly, the proteomic profile of CAA type 1 exhibited a substantial overlap with CADASIL, featuring a notable accumulation of HTRA1. However, it presented a distinct cerebrovascular signature compared to AD brains without vascular pathologies. In contrast, our findings indicate that protein alterations in all AD cases showed consistent directionality, irrespective of CAA severity and lacked HTRA1-dependent changes in the vascular proteome. This suggests the potential existence of divergent mechanisms in the development of different forms of CAA. For example, CAA type 1 is predominantly observed in capillaries whereas the more common type 2 of CAA form demonstrates amyloid in leptomeningeal and penetrating arteries and arterioles, without capillary involvement.

A recent large-scale multi-omics analysis of over 500 cases across multiple sample collections and brain regions uncovered a ‘Matrisome’ module (M42) that dramatically increased in AD on the protein level but was not observed in the RNA network^29^. Our results affirm the reproducibility of prior findings with a consistent observation of matrisome proteins in M89 in our study showing significantly elevated levels in AD compared to cognitively normal control and non-AD tauopathy brain tissues. This pattern was evident in both bulk and vascular proteomes. Several members of the matrisome module, including SMOC1, MDK, NTN1 have been previously shown to bind Aβ and co-localize with amyloid plaques in AD brains^29,49^ and in a mouse model of AD amyloidosis^31,33^. In the current study, beyond the core proteins that are central to the module’s function, we have identified novel markers-SMOC2, ITM2C, HGF, CHADL, and others- that were not discernible at the bulk level using similar database search algorithms. Significantly, we have observed distinctly elevated levels of these proteins in response to increased amyloid deposition in the human cerebrovasculature. SMOC1 (secreted modular calcium-binding protein 1), exhibiting binding affinity to heparin and heparan sulfates via glycosaminoglycan-binding-like motifs, plays a crucial role in amyloid deposition, cell adhesion, and angiogenesis^50^. In addition, recent systems-based AD biomarker studies showcased SMOC1 as a reliable CSF biomarker reflective of pathological processes occurring in AD brain^51-53^. Given the high homology between SMOC1 and SMOC2, the functions attributed to SMOC1 can likely be extended to less characterized SMOC2^54^. Of note, in our study SMOC2 has not shown association with amyloid plaques, but rather high specificity to vascular amyloid suggesting that it may play an important role in CAA formation. Although not extensively studied, other M89 members may influence AD-associated pathology through different mechanisms of action, including APP processing. ITM2C (Integral membrane protein 2C) also known as BRI3 has been shown to interact with APP and act as a negative regulator of Aβ production^55^. Its homolog BRI2 (ITM2B) is known to cause cerebral amyloid angiopathy in familial British and Danish dementias^56^. In contrast, SFRP1 (secreted-frizzled-related protein 1) interferes with anti-amyloidogenic processing of APP via metalloprotease ADAM10 leading to generation of toxic Aβ products^57^. Meanwhile, SLIT2 (Slit guidance ligand 2) and HGF (hepatocyte growth factor) have been implicated in angiogenesis and vascular permeability by possibly regulating BBB integrity and endothelial function^58-61^. Future mechanistic studies delineating how these proteins influence cerebrovascular health and deposition of amyloid within the walls of blood vessels may shed light on AD pathophysiology and facilitate selecting the protein targets for disease-modifying therapies. This may potentially impact transition from the biochemical to the cellular phase of AD^62^, and specifically opening up therapeutic possibilities targeting discrete pathophysiology of AD vasculature and its ECM affecting the BBB and glymphatic function.

The allelic variation in apolipoprotein E (*APOE*) has shown the strongest susceptibility for sporadic AD and CAA, with the presence of the *APOE e4* allele increasing risk in a dose dependent manner^41,63^. APOE4 accelerates breakdown of the BBB^64,65^ and increases risk for cognitive impairment during normal aging^65^. Furthermore, it has been reported that APOE4 is associated with disruption of the perivascular drainage pathway, an effect mediated by APOE-related alterations of basement membrane proteins^66,67^. In the present study, we found an inverse relationship between *APOE4* genotype and five protein modules of which M28 and M90 have been closely associated with SMC biology. Immunohistochemical surveying of microvascular changes in AD has shown significant reduction of smooth muscle actin in *APOE4* carriers compared to *APOE3* counterparts^68^. This was further corroborated by *in vitro* and *in vivo* data demonstrating the effect of APOE on SMC proliferation and migration^69,70^. Despite this evidence of direct impact of APOE on SMC function, we cannot rule out the possibility of other contributing mechanisms. Mounting evidence has demonstrated the strong association between *APOE4* genotype and CAA development in elderly individuals. In the late stages of CAA, the vessel walls are replaced by amyloid, leading to gradual SMCs degeneration and increased permeability suggesting an indirect effect of APOE on vascular cell alterations. Thus, the exact connection between APOE genotype and SMC function remains to be determined. Moreover, given the small number of *APOE4* carriers in our study, further examination of the effect of APOE on the cerebrovascular proteome is needed.

Characterizing the proteomic landscape of cerebrovasculature in tauopathies where amyloid deposition is not present has been largely limited. However, recent evidence for an association between tau pathology and vascular deficits has emerged. Cerebrovascular alterations have been directly linked to early Braak tau pathology that culminate in CAA-independent loss of SMCs and accompanying perivascular deposition of tau^71^. Moreover, in primary tauopathies the areas affected by abundant tau pathology show morphological abnormalities of the microvasculature, including thinning, tortuosity, and vascular fragmentation^72^. Aligned with this observation, diminished cerebral perfusion and white matter hyperintensities (WMH) have been significantly correlated with tau pathology and progression^73-75^. These results have bolstered a direct interplay between cerebrovascular changes and tau pathology. In our study, comparative proteomic examination of PSP cerebrovasculature found a significant increase in protein constituents of M47 and M87 strongly representative of blood coagulation and erythrocyte metabolism, respectively. Notably, despite less prominent tau pathology in frontal cortex in PSP cases, elevation in PSP of these module members compared to AD, suggests that other mechanisms may also mediate tauopathy’s effect on the brain vasculature.

Current protein biomarkers that are reliably measured in CSF of AD patients and are often used for diagnostic purposes, include Aβ_1-42_, total tau, and p-tau^46-48^. Although these CSF markers detect the pathological hallmarks of AD, they represent only a small portion of all pathophysiological processes that occur in the diseased brain. Vascular deficits appear in the early stages of AD and may precede the onset of neurodegenerative changes in the brain^39^. However, the lack of protein biomarkers reflecting diverse processes linked to brain vasculature limit development of diagnostic and therapeutic strategies. In the current study, we focused on cross-biofluid deep proteomic examination of overlap between heparin-enriched plasma and CSF from control and AD individuals and the cerebrovascular protein network. We identified a significant number of proteins that showed a congruent directionality of change between biofluid and brain proteomes. Among those, the most striking effect was observed for M89 module members, including SMOC1 and SMOC2 with their consistent increase in all three proteomes. These observations coupled with their high expression in Aβ-laden vessels make SMOC1 and SMOC2 promising biomarkers for vascular pathologies and potential surrogate markers of CAA. Our findings that matrisome-associated proteins may serve as key biomarkers have been recently corroborated by other studies from multiple patient cohorts showing that the levels of SMOC1 and other matrisome proteins are easily detectable and highly altered in AD CSF and plasma samples, even decades before the first AD symptoms appear^52,76,77^. Our results further showcase the divergent trends of multiple proteins in CSF and plasma that map to vascular specific modules. Specifically, extracellular matrix proteins (M11 module), including biglycan (BGN) showed a dramatic increase in AD plasma samples with a concomitant significant reduction in the brain and CSF. Although the exact mechanism of the discordant directions of change requires further investigation, vascular alterations and injury in response to changes of the expression of ECM proteins may be responsible. This is supported by previous studies showing that BGN regulates SMC function, potentially contributing to vascular deficits^78^. The discrepancy between cerebrovascular M11 and bulk brain is also notable as a potential disease specific signature representing differential change in vascular ECM distinct from parenchyma of bulk brain tissue.

The expanding framework of AD pathophysiology has necessitated the use of novel approaches for proteomic investigations. Employing deep proteomic profiling of brain vasculature in the context of AD holds great promise in identifying key drivers of cerebrovascular pathologies that may serve as therapeutic targets and biomarkers of disease progression.

## Data Availability

All data produced are available online at https://www.synapse.org/vascular

## Figure legends

**Supplemental Figure 1.**
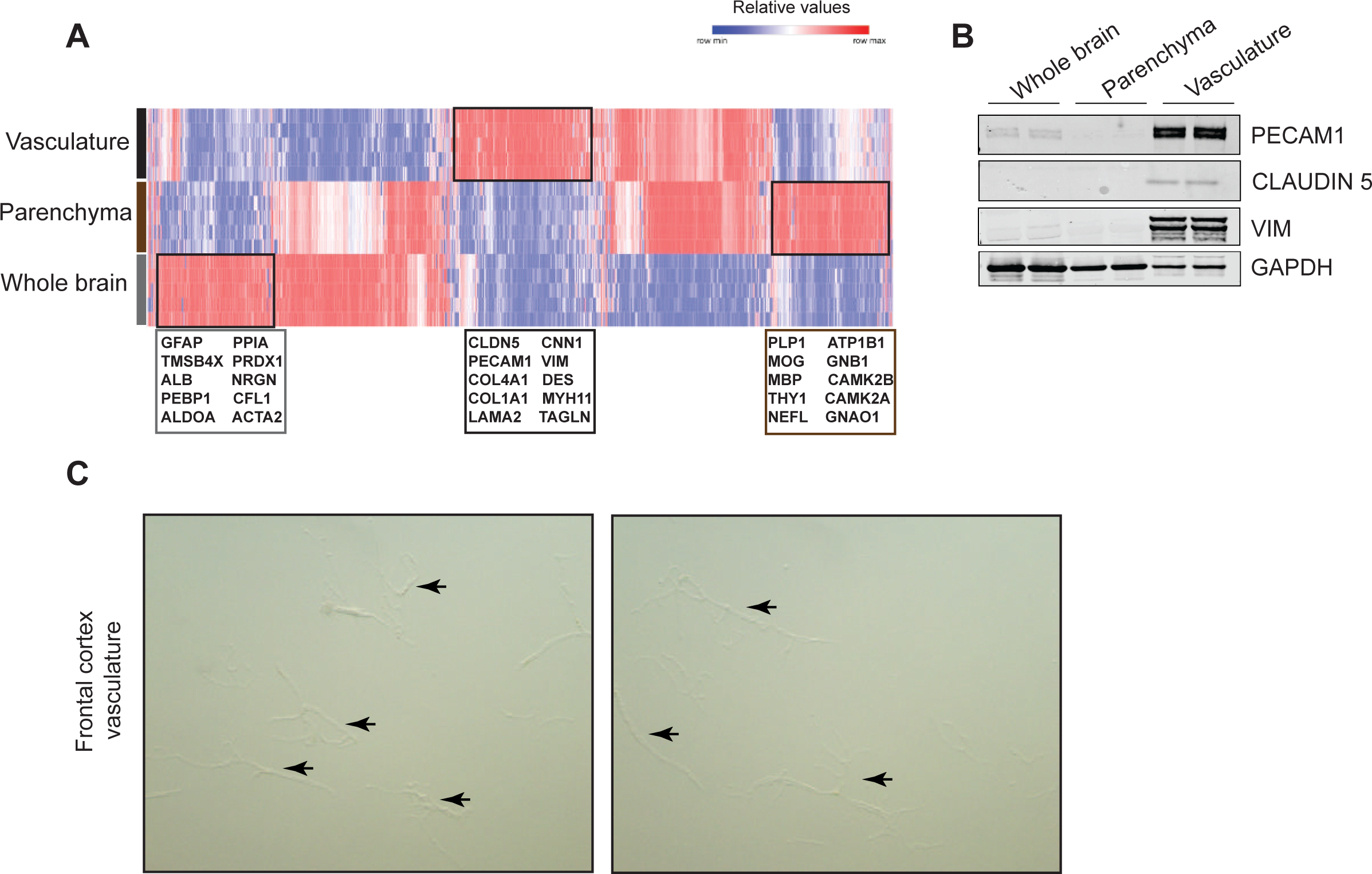
Establishment of the method of cerebrovascular isolation. (**A**) Label free mass spectrometry analysis of proteins enriched in whole brain, parenchyma, and isolated cerebrovasculature. Red indicates proteins increased and blue indicates proteins decreased in each brain fraction. (**B**) Western blot analysis shows enrichment of vascular-specific proteins in the vascular fractions compared to whole brain and parenchyma. (**C**) Microscopy images showing the purity of isolated vasculature (10x magnification).

**Supplemental Figure 2.**
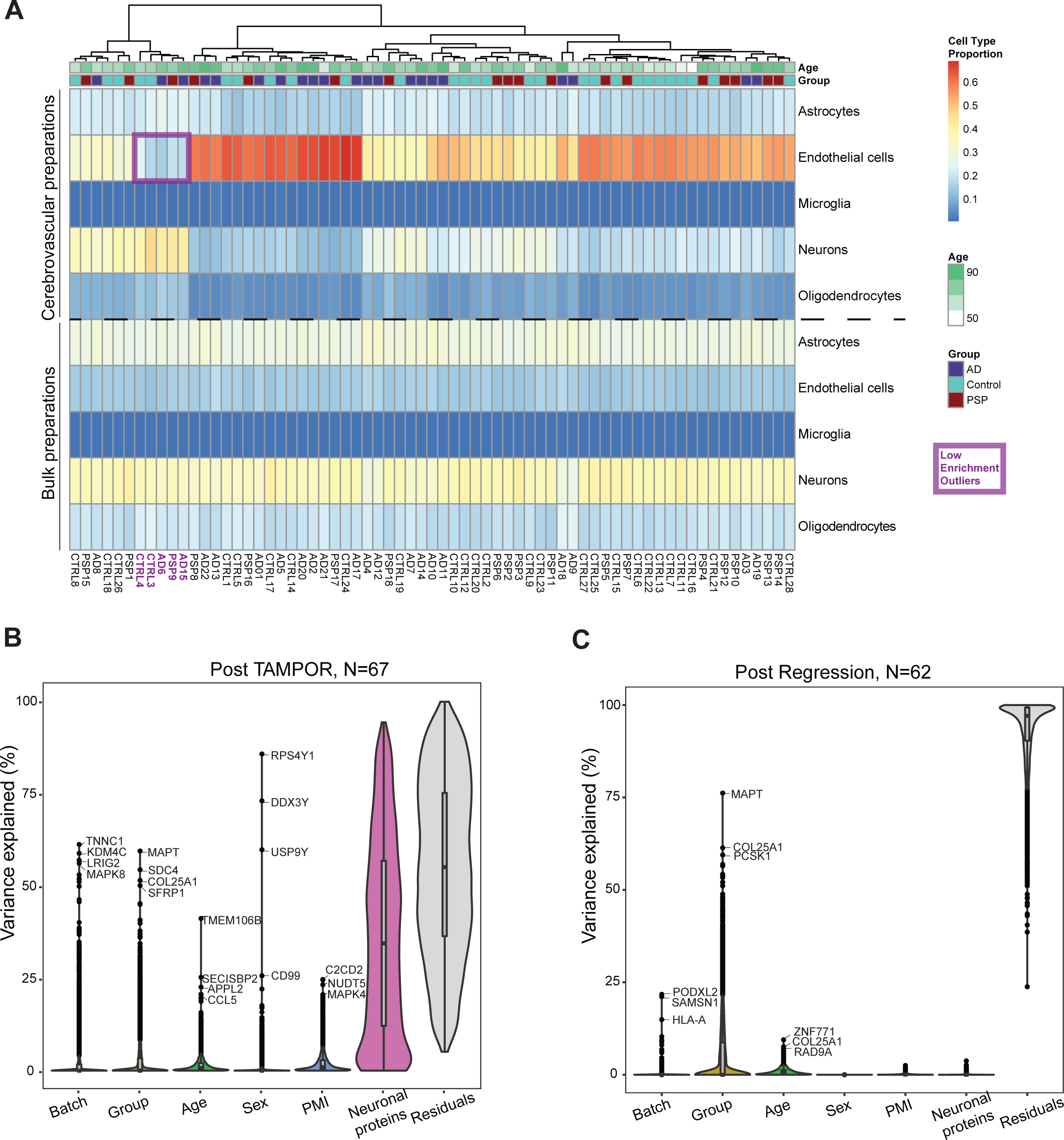
Cell type proportions in the vascular and bulk proteomes. (**A**) Heatmap showing cell type proportions in the vascular proteome (above dashed line) and bulk proteome (bottom). Proportions of the five cell type total one, and the five cell types are defined via reprocessing of Darmanis^79^, Nowakowski^80^, and Zhong single-cell datasets as curated with the EnsDeconv R package, which calculated cell type proportions by ensemble deconvolution of each proteome^81^. Dark red and orange indicate high enrichment of endothelial proteins to proportions above 70 percent, and dark blue indicates the lowest enrichment of cell-specific proteins in the preparations. The purple box outlines a cluster of outliers in the vascular preparations with very low enrichment of endothelial markers. (**B**) Variance partition plots were used to visualize the percent variance of each protein in the data set co-varying with batch, group, age, sex, postmortem interval (PMI), and neuronal proportions across case samples. (**C**) Following outlier removal, the matrix was subjected to the bootstrap regression to remove the variance due to age, sex, PMI, and neuronal cell type proportions.

**Supplemental Figure 3.**
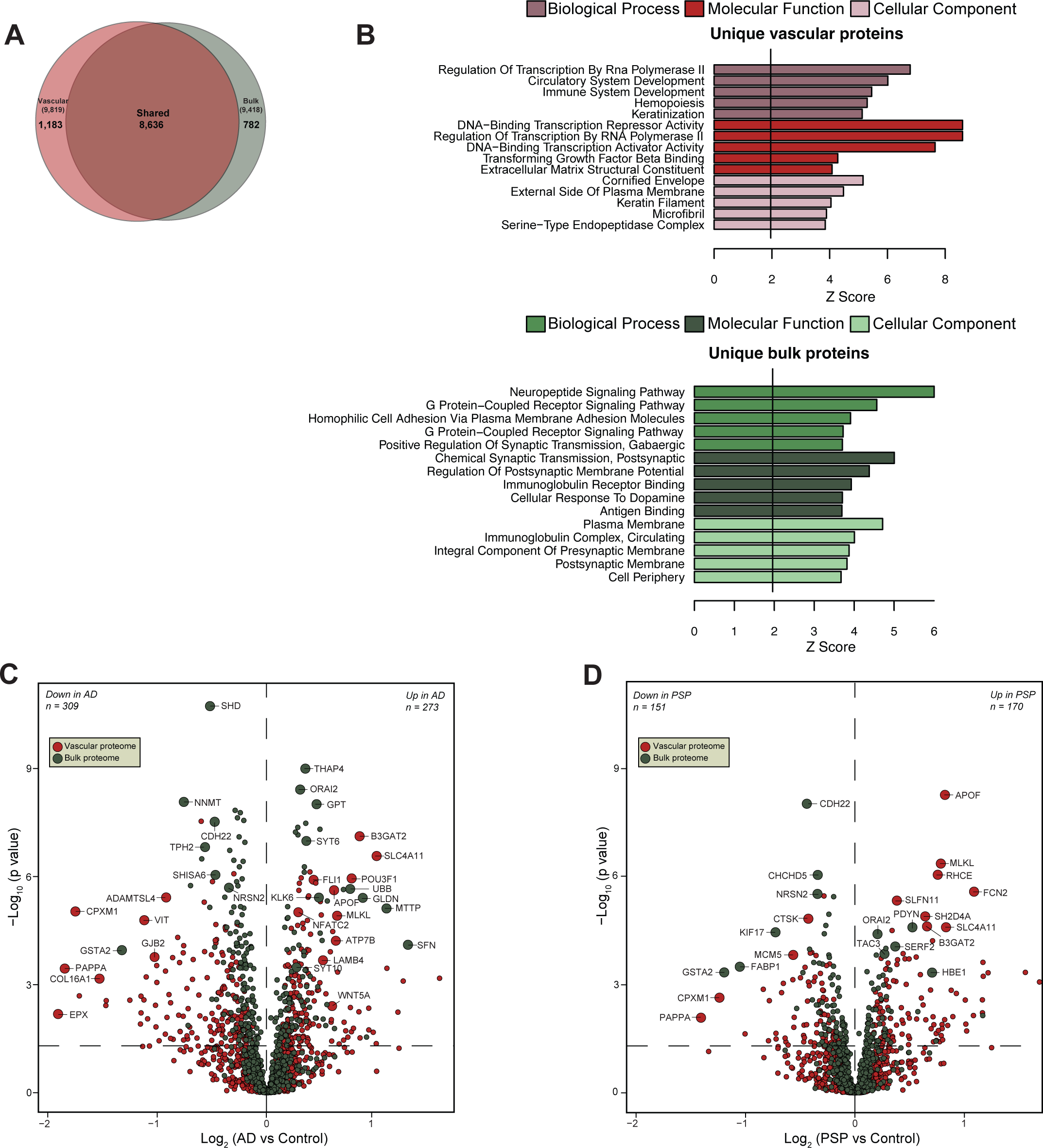
Differentially abundant proteins unique to vascular and bulk proteomes reveals specific discriminators. **(A)** Venn diagram of shared and unique proteins discovered in vascular and bulk fractions; counts shown are for unique gene products, in distinction from total protein isoforms. (**B**) GO terms enrichment analysis was used to represent biological processes, molecular function, and cellular component for unique vascular or bulk proteins. **(C**-**D)** Proteins unique to either vascular fraction or bulk proteome were input for statistics underlying volcano plots displaying differential abundance of 582 proteins between Control and AD (**C**) or 321 proteins between Control and PSP (**D**). The x axis shows the log_2_ fold change, while the y axis represents -log_10_ statistical *p* value calculated for all proteins between pairwise vascular and bulk group comparisons in AD (**C**) and PSP (**D**). One-way ANOVA was performed followed by Tukey’s post-hoc test for each pairwise comparison vs. control and imprecise Tukey p values below 10^-8.5^ were replaced with pairwise two-sided unequal variance T-tests’ p values corrected for multiple tests by Bonferroni correction. Proteins are colored based on the proteome membership (red indicates vascular proteome and green indicates bulk proteome).

**Supplemental Figure 4.**
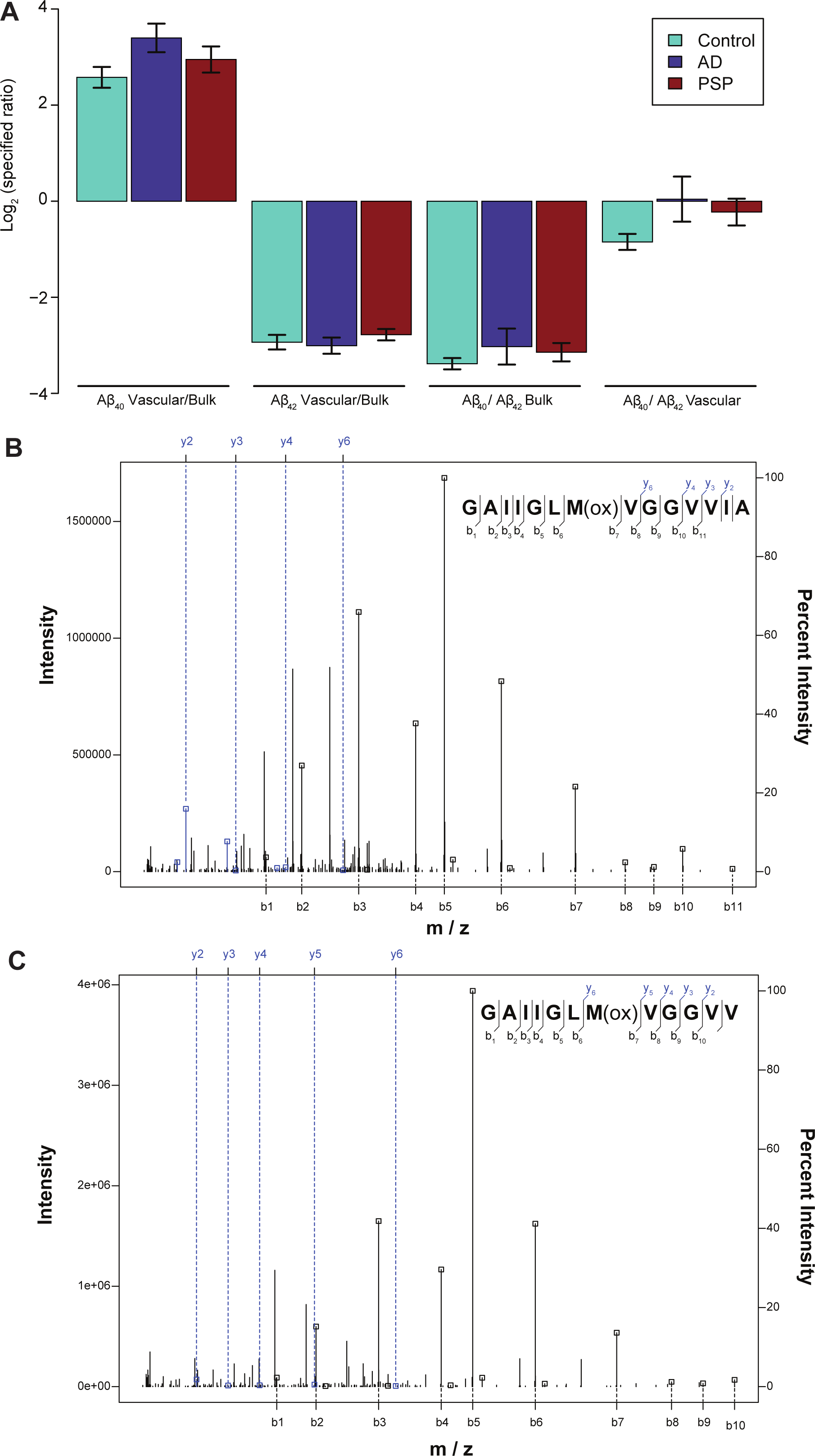
Aβ40 is predominantly found in the brain vascular fraction. (**A**) MS-quantified levels of the C-terminal tryptic Aβ40 and Aβ42 species in the vascular and bulk fractions. (**B**) MS/MS spectrum of C-terminal tryptic peptide of Aβ42 with annotation of matched B and Y ions as indicated. (**C**) MS/MS spectrum of the cognate peptide for Aβ40.

**Supplemental Figure 5.**
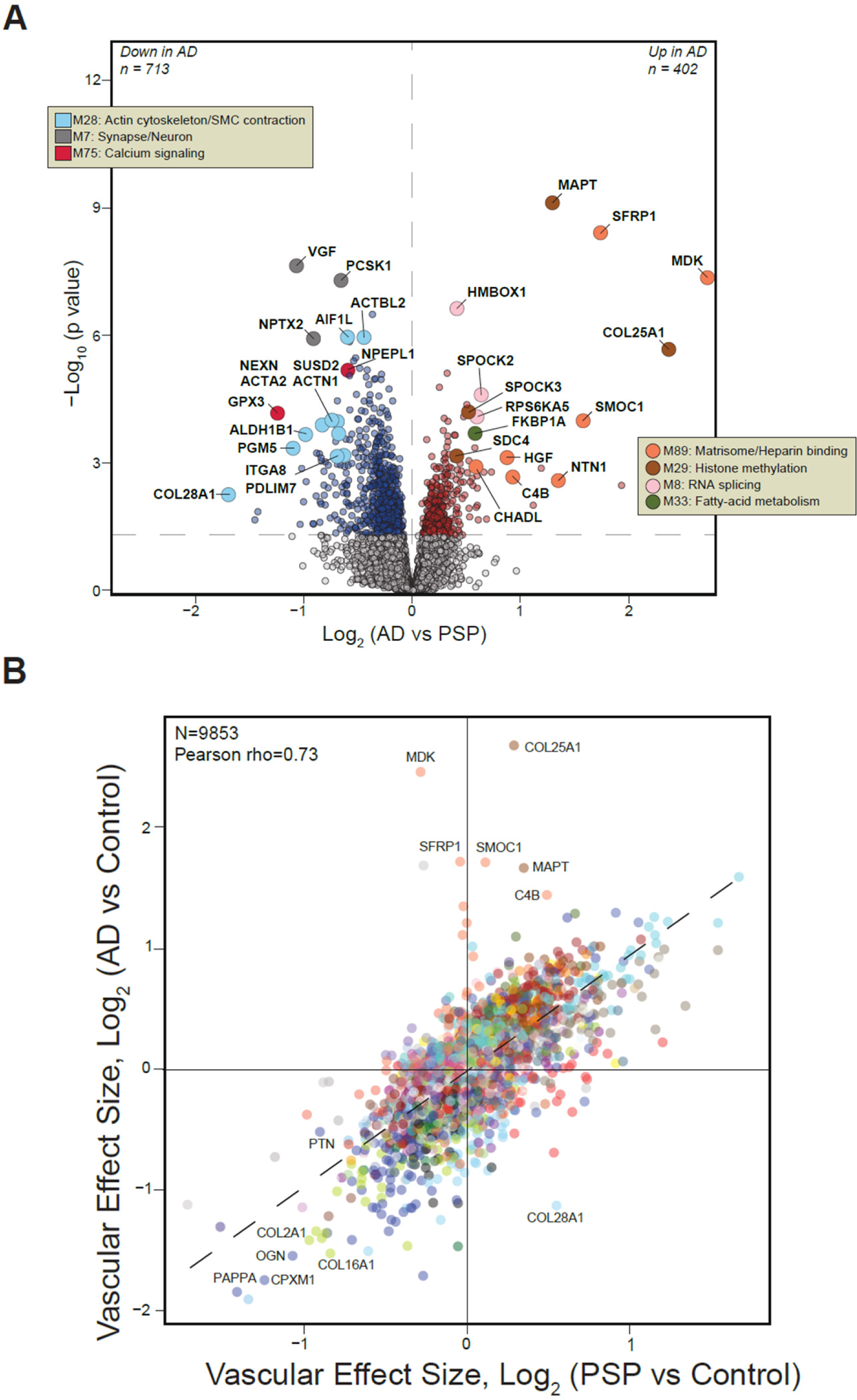
Amyloid-associated changes in cerebrovasculature drive distinct proteomic signatures in AD and PSP. **(A)** Volcano plot showing the log_2_ fold change (x axis) and -log_10_ one-way ANOVA with Tukey *p* value (y axis) for differentially changed proteins between pairwise AD and PSP comparison. Tukey *p* values below 10^-8.5^ were recalculated as Bonferroni-corrected two-tailed unequal variance t-test *p* values. Proteins are shaded based on their module membership colors. **(B)** Scatter plot of the cerebrovascular proteome log_2_ effect size of the PSP group compared to unimpaired healthy Controls (x) versus the log_2_ effect size of the AD group compared to Controls (y). Highlighted proteins showcased disease-specific signatures altered in AD and PSP.

**Supplemental Table S1. Sample characteristics - vascular and bulk proteomes.**

**Supplemental Table S2. Cell type-specific marker list from snRNA-seq.**

**Supplemental Table S3. Protein differential abundance analysis of cerebrovasculature (ANOVA).**

**Supplemental Table S4. Protein differential abundance analysis of bulk tissue (ANOVA).**

**Supplemental Table S5. Cerebrovascular WGCNA module assignment.**

**Supplemental Table S6. Gene ontology terms for cerebrovascular protein modules.**

**Supplemental Table S7. The correlation analysis of cerebrovascular protein modules.**

**Supplemental Table S8. MAGMA GWAS significance for genes associated with AD.**

**Supplemental Table S9. MAGMA GWAS significance for genes associated with PSP.**

**Supplemental Table S10. Sample characteristics - plasma proteome.**

**Supplemental Table S11. Sample characteristics - CSF proteome.**

**Supplemental Table S12. Protein differential abundance of plasma (ANOVA).**

**Supplemental Table S13. Protein differential abundance of CSF (ANOVA).**

**Supplemental Table S14. Protein overlap between plasma and cerebrovascular datasets (FET analysis).**

**Supplemental Table S15. Protein overlap between CSF and cerebrovascular datasets (FET analysis).**

## Methods

### Brain vasculature isolation from postmortem human brain and protein digestion

Postmortem frozen frontal cortex tissue was obtained from the University of Pennsylvania Brain Bank using appropriate de-identification and under proper institutional review board protocols. Cohort characteristics are presented in Supplementary Table 1. Frozen brain tissue (200-300 mg) was homogenized in ice-cold HBSS buffer with 0.1%BSA and 1% dextran (from *Leuconostoc mesenteroides*, Sigma D1537) in a glass Dounce homogenizer, as previously described. Briefly, an equal volume of 31% dextran was added to the homogenate and centrifuged at 8000 x g at 4C for 30 minutes leading to separation of myelin/parenchymal material (top layer) from vasculature (pellet). After a series of washes, the vascular-enriched pellet was resuspended in HBSS with 0.1% BSA and the vessels were captured on 40um cell strainer and then collected in a new 50 mL falcon tube. Finally, the vessels were centrifuged followed by washes with ice-cold PBS. The vessel-enriched pellet was then lysed in 8M urea lysis buffer (8M urea, 10mM Tris, 100mM NaH2PO4, pH 8.5) with HALT protease and phosphatase inhibitor cocktail (Thermo Fisher Scientific). The lysates were transferred to new Eppendorf LoBind tubes and sonicated for three cycles consisting of 5 s of active sonication at 20% amplitude, followed by 15 s on ice. Samples were then centrifuged for 5min at 15,000g and the supernatant transferred to a new tube. Protein concentration was determined by bicinchoninic acid assay (Pierce). For protein digestion, 40 μg of each sample was aliquoted, and volumes were normalized with additional lysis buffer. Samples were reduced with 1mM dithiothreitol at room temperature for 30min, followed by 5mM iodoacetamide alkylation in the dark for another 30min. Lysyl endopeptidase (Wako) at 1:100 (wt/wt) was added, and digestion was allowed to proceed overnight. Samples were then seven-fold diluted with 50mM ammonium bicarbonate. Trypsin (Promega) was then added at 1:50 (wt/wt), and digestion was carried out for another 16h. The peptide solutions were acidified to a final concentration of 1% (vol/vol) formic acid (FA) and 0.1% (vol/vol) trifluoroacetic acid (TFA) and de-salted with a 30-mg HLB column (Oasis). Each HLB column was first rinsed with 1ml of methanol, washed with 1ml of 50% (vol/vol) acetonitrile (ACN) and equilibrated with 2× 1ml of 0.1% (vol/vol) TFA. The samples were then loaded onto the column and washed with 2× 1ml of 0.1% (vol/vol) TFA. Elution was performed with 2 volumes of 0.5ml of 50% (vol/vol) ACN. An equal amount of peptide from each sample was aliquoted and pooled as the pooled global internal standard (GIS), which was split and labeled in each TMT batch as described below. Procedures for bulk tissue homogenization was performed, as previously described.

### Western blot analysis

To confirm enrichment of vascular proteins in the isolated cerebrovasculature, 10μg of protein lysates from whole brain, parenchyma, and isolated vessels were resolved on a 4–12% Bris-Tris gel and transferred onto a nitrocellulose membrane. Subsequently, the membranes were incubated over-night with anti-PECAM1, anti-CLDN5, anti-VIM, AND anti-GAPDH diluted in Start Block. After washes in TBS-T, membranes were incubated with secondary antibodies (1:10 K) and imaged using Odyssey Infrared Imaging System (LI-COR Biosciences). GAPDH signal was used for normalization.

### Sample Processing

Samples were homogenized in 8 M urea lysis buffer (8 M urea, 10 mM Tris, 100 mM NaH2PO4, pH 8.5) with HALT protease and phosphatase inhibitor cocktail (ThermoFisher) using a Bullet Blender (NextAdvance). Each Rino sample tube (NextAdvance) was supplemented with ∼100 μL of stainless-steel beads (0.9 to 2.0 mm blend, NextAdvance) and 300 μL of lysis buffer. Tissues were added immediately after excision and homogenized with bullet blender at 4 °C with 2 full 5 min cycles. The lysates were transferred to new Eppendorf Lobind tubes and sonicated for 3 cycles consisting of 5 s of active sonication at 30% amplitude, followed by 15 s on ice. Samples were then centrifuged for 5 min at 15,000 x g and the supernatant transferred to a new tube. Protein concentration was determined by bicinchoninic acid (BCA) assay (Pierce). For protein digestion, 40 μg of each sample was aliquoted and volumes normalized with additional lysis buffer. Samples were reduced with 5 mM dithiothreitol (DTT) at room temperature for 30 min, followed by 10 mM iodoacetamide (IAA) alkylation in the dark for another 30 min. Lysyl endopeptidase (Wako) at 1:25 (w/w) was added, and digestion allowed to proceed overnight. Samples were then 7-fold diluted with 50 mM ammonium bicarbonate. Trypsin (Promega) was then added at 1:25 (w/w) and digestion proceeded overnight. The peptide solutions were acidified to a final concentration of 1% (vol/vol) formic acid (FA) and 0.1% (vol/vol) trifluoroacetic acid (TFA) and desalted with a 30 mg HLB column (Oasis). Each HLB column was first rinsed with 1 mL of methanol, washed with 1 mL 50% (vol/vol) acetonitrile (ACN), and equilibrated with 2×1 mL 0.1% (vol/vol) TFA. The samples were then loaded onto the column and washed with 2×1 mL 0.1% (vol/vol) TFA. Elution was performed with 2 volumes of 0.5 mL 50% (vol/vol) ACN.

### Tandem Mass Tag (TMT) Labeling

TMT peptide labeling was performed, as previously described^28,82^. Peptides were reconstituted in 100ul of 100mM triethyl ammonium bicarbonate (TEAB) and labeling performed as previously described using TMTPro isobaric tags (Thermofisher Scientific, A44520). Briefly, the TMT labeling reagents were equilibrated to room temperature, and anhydrous ACN (200 μL) was added to each reagent channel. Each channel was gently vortexed for 5 min, and then 20 μL from each TMT channel was transferred to the peptide solutions and allowed to incubate for 1 h at room temperature. The reaction was quenched with 5% (vol/vol) hydroxylamine (5 μl) (Pierce). All 16 channels were then combined and dried by SpeedVac (LabConco) to approximately 100 μL and diluted with 1 mL of 0.1% (vol/vol) TFA, then acidified to a final concentration of 1% (vol/vol) FA and 0.1% (vol/vol) TFA. Peptides were desalted with a 30 mg HLB plate (Waters). The eluates were then dried to completeness.

### High pH Fractionation

High pH fractionation of all cases was performed essentially as described^82,83^ with slight modifications. Dried samples were re-suspended in high pH loading buffer (0.07% vol/vol NH4OH, 0.045% vol/vol FA, 2% vol/vol ACN) and loaded onto a Water’s BEH (2.1mm x 150 mm with 1.7 µm beads). An Thermo Vanquish UPLC system was used to carry out the fractionation. Solvent A consisted of 0.0175% (vol/vol) NH4OH, 0.01125% (vol/vol) FA, and 2% (vol/vol) ACN; solvent B consisted of 0.0175% (vol/vol) NH4OH, 0.01125% (vol/vol) FA, and 90% (vol/vol) ACN. The sample elution was performed over a 25 min gradient with a flow rate of 0.6 mL/min with a gradient from 0 to 50% B. A total of 96 individual equal volume fractions were collected across the gradient and dried to completeness using a vacuum centrifugation.

### Liquid Chromatography Tandem Mass Spectrometry

All samples (∼1ug for each fraction) were loaded and eluted using Dionex Ultimate 3000 RSLCnano (Thermofisher Scientific) an in-house packed 15 cm, 150 μm i.d. capillary column with 1.9 μm Reprosil-Pur C18 beads (Dr. Maisch, Ammerbuch, Germany) using a 27 min gradient. Mass spectrometry was performed with a high-field asymmetric waveform ion mobility spectrometry (FAIMS) Pro equipped Orbitrap Eclipse (Thermo) in positive ion mode using data-dependent acquisition with 1.5 second top speed cycles. Each cycle consisted of one full MS scan followed by as many MS/MS events that could fit within the given 1.5 second cycle time limit. MS scans were collected at a resolution of 60,000 (410-1600 m/z range, 4×10^5 AGC, 50 ms maximum ion injection time, FAIMS compensation voltage of -45 and -65). All higher energy collision-induced dissociation (HCD) MS/MS spectra were acquired at a resolution of 30,000 (0.7 m/z isolation width, 35% collision energy, 1.25×10^5 AGC target, 54 ms maximum ion time, TurboTMT on). Dynamic exclusion was set to exclude previously sequenced peaks for 20 seconds within a 10-ppm isolation window.

### Data Processing Protocol

All raw files were searched using Thermo’s Proteome Discoverer suite (version 2.4.1.15) with Sequest HT. The spectra were searched against a human uniprot database downloaded Feb 2019 (20342 target sequences). Search parameters included 20ppm precursor mass window, 0.05 Da product mass window, dynamic modifications methione (+15.995 Da), deamidated asparagine and glutamine (+0.984 Da), phosphorylated serine, threonine and tyrosine (+79.966 Da), and static modifications for carbamidomethyl cysteines(+57.021 Da) and N-terminal and Lysine-tagged TMT (+304.207 Da). Percolator was used filter PSMs to 0.1%. Peptides were group using strict parsimony and only razor and unique peptides were used for protein level quantitation. Reporter ions were quantified from MS2 scans using an integration tolerance of 20 ppm with the most confident centroid setting. Only unique and razor (i.e., parsimonious) peptides were considered for quantification.

### Batch correction of proteomics

Four batches of TMT 18-plex each with one global internal standard (GIS, all sample mixure), and the fourth batch with a second GIS sample, were initially corrected for batch using TAMPOR, an algorithm we developed to leverage the ratio of samples over GIS within and across batches, stabilizing and minimizing variance due to batch iteratively median-centering both proteins and samples within the two-dimensional proteomic abundance matrix, as previously described^84^. Using the option useAllNonGIS=FALSE, and with only one GIS present in three of the batches, the correction converged immediately, in 2 iterations. Code for TAMPOR as an R function is available from https://www.github.com/edammer/TAMPOR.

### Cell type proportion estimation, enrichment outlier detection, and regression of neuronal variance in vascular proteome

Cell type proportions for neurons, endothelial cells, microglia, oligodendrocytes, and astrocytes were estimated using an ensemble deconvolution approach which extracts cell type markers and marker profiles from single cell RNA-Seq (scRNA) data which had previously been clustered into the 5 desired cell types ^80,85^; these markers and/or marker profiles are then applied to deconvolute the bulk proteomics via several established algorithms like DCQ, ICeDT, FARDEEP, hspe, EPIC, or CIBERSORT, on both log- and linear transformations of the bulk abundance data (in this case, the vascular preparations from brain, as well as the paired bulk tissue), and with 5 different statistical methods for determining markers of each cell type cluster in the scRNA data (t, wilcox, combined, none, regression). The ensemble proportions are calculated from all combinations of the above variations on estimation, thereby providing more robust and confident estimates of proportions not reliant on any one method’s bias^86^. Following cell type proportion estimation, 5 samples of the 67 were found to be lacking enrichment in endothelial cells, so that these samples were considered as outliers and not carried forward in analysis. The remaining 62 non-GIS sample proteomic abundances were subjected to nonparametric bootstrap regression with a model including neuronal proportion, age, sex, PMI, and TMT batch. Each modeled trait value times the median estimated coefficient of variation from 1000 iterations of fitting for each protein was subtracted from the log_2_(abundance) matrix. AD and PSP diagnosis was also included in the model but not subtracted, thereby protecting diagnosis-specific differences in the regression of each protein. VariancePartition violin plots were generated to gauge the success of each data cleanup step, by visualizing the percetn variance explained by traits—from the starting data to the abundance matrix following TAMPOR, to that following regression.

### WGCNA definition of vascular network modules, and module preservation in bulk proteomics

A weighted protein co-expression network for the regressed cerebrovasculature log_2_(abundance) matrix of 62 samples with n=9,853 proteins having less than 50 percent missing values was built using R v4.2.3 WGCNA v1.72-1 blockwiseModules() function with the following settings. Power was 9, deepSplit was 4, minimum module size was 20, merge cut height was 0.07, TOMDenom “mean,” bicor correlation for adjacency/TOM calculation, a signed network type, partitioning about medioids staging (pamStage), pamRespectsDendro, a reassignment threshold p value of 0.05, and a maximum block size larger than the total number of proteins, i.e., built in a single block. Following blockwiseModules definition of the 93 modules, the table of kMEs calculated using bicor, and module membership based on this was iteratively checked to reassign module members subject to these rules: (1) maximum difference from the highest kME of the module to which a protein could be assigned is 0.10 (2) module membership must be assigned if any kME is greater than 0.30, and (3) must be unassigned (grey) if all kMEs are less than 0.30. The rules were met within 5 iterations out of 30 possibly allowed.

Vascular protein network module preservation in paired bulk cortical proteome was performed using the R v4.2.3 WGCNA v1.72-1 modulePreservation function over 500 permutations with the 62-sample vascular protein abundance matrix and module assignments as template network, and the paired 62 sample bulk cortex protein abundance matrix as target.

### Statistics, enriched ontology determination, and visualization

Statistics and graphical visualization were performed in R v4.2.2. Volcano plots used the custom plotvolc function available from https://www.github.com/edammer/parANOVA, which also provides the parANOVA.dex function for fast parallel calculation of one-way ANOVA+Tukey-corrected pairwise comparison p values, with fallback to Bonferroni correction for Tukey p values < 1e-8.5. ComplexHeatmap package pheatmap generated the segemented heatmaps in Figure 5C. Permutation-based assessment of normalized enrichment scores for MAGMA GWAS gene-level nominally significant hits either as an ensemble of MAGMA-processed summary statistics from three AD GWAS studies^41,87,88^ or the available PSP GWAS^89^ was performed using the MAGMA-SPA function available from https://www.github.com/edammer/MAGMA.SPA. One-tailed Fisher’s exact test heatmaps for enrichment of gene products in modules were rendered using https://www.github.com/edammer/CellTypeFET. Ontologies enriched by module were determined using GOparallel function and supporting online resources which are available via https://www.github.com/edammer/Goparallel. Module igraphs (Figure 6A) were rendered using the buildIGraphs function available from https://www.github.com/edammer/netOps. Additional visualization layout and formatting were performed in Illustrator 2022.

### Amyloid beta quantification and ratios in bulk and vascular paired samples

For amyloid beta (Aβ) quantification across both bulk and vascular samples, raw files (96 files or fractions per batch) were searched using FragPipe (version 20.0) to ensure parsimonious protein identifications. The FragPipe pipeline relies on MSFragger v3.8 ^90,91^ for peptide identification and Philosopher (version 5.0.0; da Veiga et al. 2022) for FDR filtering and downstream processing. The peptide search was done against all canonical Human proteins downloaded from Uniprot (20,402; accessed 02/11/2019), as well as common contaminants, and all reverse sequences (decoys) (n=20,453). We also included the Aβ40 (DAEFRHDSGYEVHHQKLVFFAEDVGSNKGAIIGLMVGGVV) and Aβ42 (DAEFRHDSGYEVHHQKLVFFAEDVGSNKGAIIGLMVGGVVIA) cleavage products as independent protein entries to map the C-terminus tryptic peptides.

The workflow used in FragPipe followed parameters as listed below. Briefly, precursor mass tolerance was -20 to 20 ppm, fragment mass tolerance was set to 20 ppm, mass calibration and parameter optimization were selected, and the isotope error was set to -1/0/1/2/3. The enzyme specificity was set to strict-trypsin and up to two missed cleavages allowed. Cleavage type was fully enzymatic. Peptide length was allowed in the range from 7 to 50 and peptide mass from 200 to 5,000 Da. Variable modifications that were allowed in our search included: oxidation on methionine, N-terminal acetylation on protein, TMTpro (TMT16, +304.20715) modifications on serine, threonine and histidine, with a maximum of 3 variable modifications per peptide. Static modifications included: isobaric TMTpro (TMT16) modifications on lysine and the peptide N-terminus as well as carbamidomethylation of cysteine (+57.02146). Peptide spectral matches (PSMs) were validated using Percolator^92^ with a false discovery rate (FDR) threshold was set to 1%. Protein and peptide abundances were quantified using Philosopher. Following quantification, peptide levels were scaled by multiplying each peptide intensity by the ratio of the sum of all the reporter ion intensities of the TMT channel (sample) with the maximum summed intensity over the current channel-specific peptide summed reporter ion intensity.

Following the search, peptide-level quantitation of the full data for bulk and separately, for vascular proteomes was subjected to two-way median polish with TAMPOR using only GIS samples within batch as denominators (useAllNonGIS=FALSE), available from https://www.github.com/edammer/TAMPOR^84^, and then the batch-corrected, normalized reporter ion intensities for the C-terminal tryptic peptides of Aβ40 (GAIIGLMVGGVV) and Aβ42 (GAIIGLMVGGVVIA) in the 62 samples which were the focus of this study were extracted from the larger dataset and plotted as ratioed values for vascular/bulk and Aβ40/Aβ42.

Representative MS/MS peptide spectral matches for Aβ40 and Aβ42 C-terminal peptides (Extended Data Fig. 4) were identified in FragPipe PSM-level summary tab-separated value files and extracted and semi-automatically annotated for matching b and y ions using a custom in house R script, available from https://www.github.com/edammer/MQ1pepAnnotate. Additional formatting was performed in Illustrator 2022.

### Protein differential abundance and overrepresentation analysis between brain vascular and biofluids proteome

A detailed description of CSF collection, clinical evaluation and proteomic analysis has been previously provided^45^. Here, TMT-MS raw data from approximately 300 CSF samples collected at the Emory Goizueta Alzheimer’s Disease Research Center (ADRC) were retrieved and processed using FragPipe with search parameters as previously outlined^44,45^. Volcano plots, illustrating differential abundance, were generated using the ggplot2 package in R v.4.2.1. Pairwise differentially abundant proteins were identified through Student’s t-test, followed by Benjamini-Hochberg (BH) false discovery rate (FDR) correction. For plasma samples, a prior differential expression analysis was conducted on 109 individual samples to produce a composite p-value, considering both the significance of association and effect size (AD vs Control) for 2865 heparin-enriched plasma proteins^44^. Only individuals meeting stringent CSF biomarker criteria (tTau/Aβ1-42 ratio > 0.226 for Alzheimer’s Disease) or Montreal Cognitive Assessment (MoCA) cutoffs (Alzheimer’s Disease < 24, Control > 26) at the time of lumbar puncture were included in the analysis. All plasma and CSF proteins as well as those significantly altered in AD compared to controls (Student’s *p* < 0.05) were assessed for overrepresentation in brain vascular proteome using a one-tailed FET, and those modules with BH FDR-corrected *p* < 0.05 were considered significant. The background for this overrepresentation analysis comprised 9854 UniprotID-identified proteins from 93 co-expression protein modules.

### Immunohistochemical analysis

To validate the proteins of interest, human postmortem brain tissues were obtained from the Emory University Brain Bank from age-matched individuals diagnosed with Alzheimer’s disease, progressive supranuclear palsy, and non-demented controls. For histological analyses, paraffin-embedded frontal cortical sections were cut at 5 μm, then deparaffnized using xylene and rehydrated in a graded series of alcohols. Antigen retrieval was achieved by incubating tissue sections in distilled water for 30 min under high temperature. The brain sections were incubated with 3& hydrogen peroxidase for 5 min and blocked in goat serum for 1 hr, followed by overnight incubation with anti-SMOC1, anti-SMOC2 (generous gift of Dr. Todd Golde), anti-MDK (Invitrogen 1:250), anti-SLIT2 (Invitrogen 1:250), anti-HHIPL1 (Invitrogen 1:250), anti-ITM2C (Invitrogen 1:250). After washes, the sections were incubated with biotinylated antibody for 30 min, followed by incubation with ABC reagent (Vectastain ABC-HRP kit, Vector Laboratories). The sections were then dehydrated, counterstained with hematoxylin, and coverslipped using permanent mounting media. The imaging was performed using Keyence microscope.

### Availability of data and materials

Raw mass spectrometry data and pre- and post-processed plasma protein abundance data and case traits related to this manuscript are available at https://www.synapse.org/vascular. The AMP-AD Knowledge Portal is a platform for accessing data, analyses and tools generated by the AMP-AD Target Discovery Program and other programs supported by the National Institute on Aging to enable open-science practices and accelerate translational learning. The data, analyses and tools are shared early in the research cycle without a publication embargo on secondary use. Data are available for general research use according to the following requirements for data access and data attribution (https://adknowledgeportal.synapse.org/#/DataAccess/Instructions).

## Funding

This study was supported by the following National Institutes of Health (NIH) funding mechanisms [U01AG061357 (A.I.L and N.T.S), RF1AG062181 (N.T.S.) and P30AG066511 (A.I.L)], the Foundations for the National Institute of Health AMP-AD 2.0 grant and a grant from the Alzheimer’s Association (ABA-22-974673).

---

| Sample Size | Control<br>N= 28 | AD<br>N= 21 | PSP<br>N= 18 |
| --- | --- | --- | --- |
| **Characteristics** |  |  |  |
| **Age, years** ± SD | 65.8 ± 8.0 | 78.8 ± 10.5 | 76.3 ± 5.5 |
| **Sex, N Males** | 15 | 9 | 11 |
| **N Females** | 13 | 12 | 7 |
| **PMI, years** ± SD | 14.6 ± 5.3 | 12.2 ± 5.4 | 11.6 ± 6.2 |
| **Amyloid burden, CERAD 0-1** | 27 | 0 | 17 |
| **CERAD 2-3** | 1 | 21 | 1 |
| **Tau burden, Braak 0-I** | 28 | 0 | 9 |
| **Braak, II-III** | 0 | 21 | 9 |
| **CAA score, 0 - no CAA** | 25 | 2 | 14 |
| **1 - mild CAA** | 2 | 7 | 3 |
| **2 - moderate CAA** | 0 | 5 | 1 |
| **3 - severe CAA** | 1 | 7 | 0 |
| **APOE genotype, E44** | 1 | 6 | 0 |
| **E34** | 5 | 4 | 2 |
| **E33** | 19 | 11 | 9 |
| **E24** | 2 | 0 | 0 |
| **E23** | 1 | 0 | 5 |
| **Abbreviations:** Ctrl, Control; AD, Alzheimer's disease; PSP, progressive supranuclear palsy; M, males; F, females; PMI, postmortem interval; APOE, apolipoprotein E |  |  |  |

| Sample Size | Ctrl | AD |
| --- | --- | --- |
|  | N= 141 | N= 140 |
| **Characteristics** |  |  |
| **Race**, *African American* | 18 | 10 |
| *Caucasian* | 122 | 129 |
| *Other* | 1 | 1 |
| **Age**, *years* ± *SD* | 64.7 ± 7.8 | 68.1 ± 8.4 |
| **Sex**, *N Males* | 37 | 65 |
| *N Females* | 104 | 75 |
| **APOE genotype**, *E44* | 3 | 28 |
| *E34* | 29 | 71 |
| *E33* | 75 | 38 |
| *E24* | 7 | 2 |
| *E23* | 25 | 1 |
| **Abbreviations:** Ctrl, Control; AD, Alzheimer's disease; M, males; F, females; APOE, apolipoprotein E |  |  |

## Notes

### Competing Interest Statement

The authors have declared no competing interest.

### Author Declarations

The Institutional Review Board (IRB) of the University of Pennsylvania gave ethical approval for this work.

